# Prognosis of stroke subtypes in whole population health systems data: a matched cohort study

**DOI:** 10.64898/2026.04.17.26351150

**Authors:** Alice Hosking, Matthew H Iveson, Laura Sherlock, Mome Mukherjee, Claire Grover, Beatrice Alex, Sai Parepalli, Grant Mair, Fergus Doubal, Heather Whalley, Richard Tobin, Joanna Wardlaw, Rustam Al-Shahi Salman, William N Whiteley

**Author notes:** **Corresponding author:** Dr Alice Hosking. E. A: Chancellor’s Building, 49 Little France Crescent, Edinburgh EH16 4SB.

## Abstract

**Background:** Outcome after stroke varies according to stroke subtype by location, but healthcare systems data studies do not include subtyping information. We linked natural language processing (NLP) of brain imaging reports to routinely collected data to estimate risk of death and other outcomes after stroke subtypes in a nationwide dataset.

**Methods:** We applied a previously validated NLP algorithm to all CT and MRI head scan reports in Scotland between 2010 and 2018. We linked the reports to hospital readmissions, prescriptions and death data to identify and characterize people with stroke, and to categorize into deep and cortical ischemic stroke, deep and lobar intracerebral hemorrhage (ICH), subarachnoid hemorrhage, and subdural hemorrhage. We used a matched cohort design, and age- and sex-matched four controls per case who never had a stroke. By subtype, we estimated rehospitalization with stroke, myocardial infarction (MI), cancer, dementia, epilepsy and death, accounting for confounders and competing risk of death.

**Results:** From 785,331 people with a head scan, we identified 64,219 with clinical stroke phenotypes (mean age 73.4yrs, 49.5% male), and subtyped 12,616 with deep ischemic stroke; 14,103 with cortical ischemic stroke; 1,814 with deep ICH; and 1,456 with lobar ICH. There was higher absolute rate of 1-year hospital readmission for lobar compared with deep ICH (4.9% [95%CI 3.9% - 6.1%] vs 3.4% [2.6% - 4.3%]), higher risk of dementia beyond 6 months after lobar ICH compared to controls than for other stroke subtypes (aHR 3.5 [2.3-5.3]); and higher risk of MI within 6 months of cortical ischemic stroke than for other stroke subtypes (aHR 4.6 [3.4-6.3]).

**Conclusions:** NLP of free-text reports linked to coded data successfully subtyped stroke at scale, and we estimated risk of clinically relevant outcomes. Future work should use free text to enable large-scale audit and epidemiology of people with stroke.

## Introduction

Outcome after stroke varies by stroke type and stroke location, but existing studies of stroke subtypes by location are limited by coding systems (“ontologies”) in healthcare data; and in cohort studies by cohort size, consent bias, and limited follow up. ^1–3^

Healthcare systems data (HSD) provide the opportunity to examine outcomes after stroke in a large number of unselected people, reducing uncertainty and increasing relevance to clinical practice. ^4–7^ However, phenotyping in HSD is constrained by coding ontologies. Health systems typically record ischemic stroke, intracerebral hemorrhage (ICH) and subarachnoid hemorrhage (SAH) with ontologies such as the 10^th^ revision of the International Classification of Diseases (ICD-10),^8,9^ but ICD-10 codes give no information about ischemic stroke location, most people with intracerebral hemorrhage do not have a stroke location recorded, and many patients are not coded as ischemic or hemorrhagic stroke.^10^ Although ICD-11 expands cerebrovascular granularity and SNOMED-CT can represent subtypes, they are not adopted widely for use in hospital coding systems.

Uncoded data (e.g., text or images) could improve phenotyping. Imaging reports are concise, clinically focused, and can be read with natural language processing (NLP). We previously developed and validated a rules-based NLP system for brain imaging reports, Edinburgh Information Extraction for Radiology reports (EdIE-R), which accurately tags MRI and CT head reports with stroke and subdural hemorrhage (SDH) types and locations.^11–13^ Demonstrating that uncoded data can improve phenotyping of stroke would be useful to healthcare and research.

In this study, we identified cortical and deep ischemic strokes, and lobar and deep ICH with NLP from free-text CT and MRI head scan reports in Scotland and linked coded HSD, and estimated risks of death and other outcomes after stroke subtypes by location. We age- and sex-matched people with stroke to controls without stroke. We examined outcomes relevant to stroke survivors: hospital readmission, cancer, epilepsy, dementia, myocardial infarction (MI) and death.

## Methods

### Data Sources

We accessed CT and MRI head scan reports in adults (≥18 years) from Scottish Medical Imaging from 1^st^ January 2010 to 31^st^ August 2018.^14^ Public Health Scotland deterministically linked imaging data with a unique patient identifier (Community Health Index number) to: general and psychiatric hospital admissions (Scottish Morbidity Record (SMR)01 and SMR04); the Scottish Cancer Registry (SMR06); community prescriptions (Prescribing Information System, from 1^st^ January 2007 to 31^st^ December 2020); and death records from NRS Scotland (from 1^st^ January 2010 to 31^st^ December 2020). Prescription data included drugs for vascular diseases, dementia and epilepsy (Supplement 2).

We tagged scan reports with an NLP algorithm developed and validated with a subset of the Scottish data used in this study. Our previous work showed that the algorithm had good precision and reasonable recall for each phenotype.^11–13^ The algorithm generated the following tags from text in the radiology report: ischemic stroke, ICH, SAH, SDH and stroke of unknown type (when a stroke was identified but not the type). Ischemic stroke and ICH were subclassified as recent or old, and cortical (for ischemic stroke), lobar (for ICH) or deep (i.e. in deep brain structures). Additional tags included atrophy and white matter hypoattenuation (on CT) or hyperintensities (on MRI) (WMH).

### Population

Almost all healthcare in Scotland is delivered by the National Health Service (NHS). People with a stroke can present either to primary care, where they may be referred on to the emergency department or for outpatient assessment; or directly to the emergency department; or be identified only at death, via autopsy. People in the emergency department may either be discharged, be admitted to hospital, or die in the emergency department. Primary data is not currently available in Scotland, and outpatient and emergency department data does not code stroke types. We defined cases as people with all the following: a stroke/SDH ICD-10 code in hospital admission and death data in the main condition/primary cause of death field (code lists in Supplement 2; a head scan within 30 days of the ICD-10 code; and no history of recorded stroke in hospital admission between Scotland from 1^st^ January 2010 to 31^st^ August 2018. We included spontaneous SDH as a spontaneous vascular event that leads to focal or global neurological symptoms. Follow-up for outcomes continued to 31^st^ December 2020.

### Stroke Cohort and Matched Cohort

We defined six stroke (or SDH) categories: ischemic stroke, ICH, SAH, SDH, stroke unspecified and non-traumatic hemorrhage unspecified; and four location subtypes: cortical and deep ischemic stroke, and lobar and deep ICH. For each case identified in hospital or death records, we compared the stroke-related ICD-10 code and NLP tag and used an algorithm to assign diagnosis where the two differed (Supplement 1, eTable 1). The algorithm prioritized common stroke types (e.g. ischemic stroke over intracerebral hemorrhage), and recent strokes over old. We then used NLP to assign location subtypes for ischemic stroke and ICH. We removed people with head trauma ICD-10 codes within seven days of ICH, or brain cancer codes within one year of ICH, as these indicate bleeding due to non-vascular pathology (brain tumor).

A matched cohort without recorded stroke/SDH was drawn from people with a record in hospital admission data, community prescription data or death data. For each case, four controls were matched on year of birth (+/− 2 years) and sex. Controls had to be alive at the event date of their matched case.

Follow-up began at admission date for the index event (for controls, the date of their matched case’s index event) and stopped at date of death or each outcome. We did not have reliable emigration data, but less than 2% of the Scottish population emigrate annually.^15^

### Outcomes

We defined death, readmission with stroke, dementia, epilepsy, dementia, cancer and MI (Supplement 2). We ascertained outcome events for stroke/SDH types and subtypes which correspond to defined pathological phenotypes: ischemic stroke, ICH, SAH and SDH. We tabulated the most common underlying causes of death for each stroke type.

Readmission with stroke was defined as hospital admission with a stroke/SDH ICD-10 code in the primary position, starting from date of discharge after the index event. For cancer, epilepsy and MI, we included relevant ICD-10 codes in the primary position in hospital admission data or underlying cause of death from the stroke date onwards. We included dementia in any position, but excluded people with events coded on the same admission as the index stroke, which likely represented pre-existing dementia. We also identified prevalent and incident epilepsy and dementia through prescriptions of anti-epileptic or dementia drugs before or after the index date; and prevalent and incident cancer cases in cancer registry data.

### Covariates

Atrial fibrillation, diabetes and hypertension were identified in hospital records with ICD-10 codes in any position, before the date of defining stroke. Additionally, hypertension and diabetes mellitus were identified with antihypertensive or diabetic medication prescriptions in the year prior to the index event (Supplement 2). We identified statin, anticoagulant and antiplatelet prescriptions in the year before the index event. We recorded the number of hospital admissions and prescriptions dispensed in the year before the index event. Additional phenotypes identified with NLP were atrophy and WMH.

### Statistical Methods

We calculated cumulative incidence of each event type, accounting for competing risk of death and censoring on death or 31^st^ December 2020, and report by sex and age group (≤70 years and >70 years). We used cause-specific Cox proportional hazard models to estimate adjusted hazard ratios (aHRs) with 95% CIs, adjusted for potential confounders (age, sex, diabetes, hypertensive, atrial fibrillation, statins, anticoagulants, antiplatelets, number of prescribed medicines in previous year, number of hospital admissions in previous year). We compared each stroke type and subtype with their control group for each outcome, except for readmission with stroke, where we compared stroke types to ischemic stroke, as those without stroke could not be readmitted. For stroke subtypes, we compared deep strokes to cortical/lobar.

As events were much more frequent in the first 6 months after stroke, the proportional hazards assumption was violated, so we split our analysis into two periods: the first 6 months after stroke, and 6 months onwards. For each outcome, we confirmed the proportional hazards assumption with visual inspection of plots of Schoenfeld residuals, and confirmed the linearity of continuous input covariates or their logarithm by inspection of plots of Martingale residuals.

Analyses were performed within the NHS Scotland Trusted Research Environment in R, version 4.4.2, using the packages survival and survminer.

### Standard Protocol Approvals, Registrations, and Patient Consents

The study was approved by the NRES Committee Northwest - Greater Manchester East ethics committee (15/NW/0719), and the Public Benefit and Privacy Panel for Health and Social Care (1516-0219) of NHS Scotland. Participant consent was not required.

### Data Availability

Data is restricted as per Public Health Scotland guidelines, as it relates to individual health records. Data can be made available within the National Safe Haven upon application and approval by Public Health Scotland and the Public Benefit and Privacy panel.

## Results

Of 5,667,954 people with any record in prescription, imaging, hospital or death data, 785,331 had a CT or MRI head scan. Of these, 79,045 had an ICD-10 stroke code recorded at any time, and 66,917 had brain imaging within 30 days of a recorded stroke (Supplement 1, eFigure 1). The median follow-up time of this population was 3.3 years (IQR 0.46 – 5.82 years) and of their matched controls was 4.9 years (IQR 3.1 years to 7.3 years).

In hospital admission and death data, 34,739 people had an ICD-10 diagnosis of ischemic stroke, 7,075 had ICH and 17,472 had unspecified stroke within 30 days of a head scan. No further subtyping was possible. We selected the brain scan closest in time to the stroke diagnosis, of which 64,178 were CT head scans, and 2,739 were MRI head scans. After using NLP, 64,219 people had recognized clinical stroke phenotypes (mean age 73.4yrs, 49.5% male). 51,339 people had ischemic stroke [12,616 (24.6%) deep, 14,103 (27.5%) cortical and 24,620 (48.0%) uncertain or multiple locations], 7,182 had ICH [1,814 (25.2%) deep, 1,456 (20.3%) lobar and 3,912 (54.5%) uncertain or multiple locations], 2,984 had SAH, 2,714 had SDH, and 2,618 unspecified stroke. Using NLP reduced the proportion of people with unspecified stroke from 26.1% to 3.4% (Supplement 1, eFigure 2).

Compared with cortical ischemic stroke, those with deep ischemic stroke were more often male (52.2% vs 49.9%), had lower prevalence of hypertension (66.6% vs 71.6%) and atrial fibrillation (12.1% vs 18.9%), and higher rates of atrophy (52.8% vs 48.6%) and WMH (68.5% vs 59.3%). Compared with lobar ICH, more of those with deep ICH were male (50.3% vs 45.1%) and fewer had taken antiplatelets (33.9% vs 40.6%) (Table 1).

**Table 1:** Baseline data of people with ischemic stroke (IS), intracerebral hemorrhage (ICH), and subtypes by location. WMH= white matter hypointesities (CT)/hyperintensities(MRI).

|  | Ischemic stroke (51,339) |  |  |  | Intracerebral hemorrhage (7182) |  |  |  |
| --- | --- | --- | --- | --- | --- | --- | --- | --- |
|  | Cortical<br>(14,103) | Deep<br>(12,616) | Cortical and<br>deep (768) | No location<br>(23,852) | Lobar<br>(1456) | Deep<br>(1814) | Lobar and<br>deep (145) | No location<br>(3767) |
| Sex (male) | 7038 (49.9%) | 6587 (52.2%) | 361 (47.0%) | 11695 (49.0%) | 656 (45.1%) | 912 (50.3%) | 61 (42.1%) | 1785 (47.4%) |
| Age (yrs, mean (SD)) | 75.8 (12.6) | 74.5 (12.9) | 74.2 (13.0) | 72.7 (13.9) | 75.2 (13.0) | 73.3 (14.3) | 75.3 (11.6) | 73.1 (14.0) |
| Atrophy (n,%) | 6854 (48.6%) | 6661 (52.8%) | 346 (45.1%) | 10,805 (45.3%) | 644(44.2%) | 804(44.3%) | 61 (42.1%) | 1386 (36.8%) |
| WMH (n,%) | 8363 (59.3%) | 8642 (68.5%) | 467 (60.8%) | 13,095 (54.9%) | 823 (56.5%) | 1050 (57.9%) | 72 (49.7%) | 1793 (47.6%) |
| Diabetes (n,%) | 1942 (13.8%) | 1680 (13.3%) | 111 (14.5%) | 2903 (12.2%) | 150 (10.3%) | 168 (9.3%) | 12 (8.3%) | 361 (9.6%) |
| Hypertension (n,%) | 10101 (71.6%) | 8402 (66.6%) | 504 (65.6%) | 15,161 (63.6%) | 902 (62.0%) | 1154 (63.6%) | 84 (57.9%) | 2298 (61.0%) |
| Atrial fibrillation (n,%) | 2671 (18.9%) | 1525 (12.1%) | 140 (18.2%) | 3108 (13.0%) | 195 (13.4%) | 227 (12.5%) | 14 (9.7%) | 439 (11.7%) |
| On statins (n,%) | 7239 (51.3%) | 5812 (46.1%) | 343 (44.7%) | 10,502 (44.0%) | 666 (45.7%) | 773 (42.6%) | 79 (48.3%) | 1621 (43.0%) |
| On anticoagulants (n,%) | 1710 (12.1%) | 1100 (8.7%) | 88 (11.5%) | 2263 (9.5%) | 217 (14.9%) | 310 (17.1%) | 21 (14.5%) | 568 (15.1%) |
| On antiplatelets (n,%) | 6729 (47.7%) | 5383 (42.7%) | 309 (40.2%) | 9196 (38.6%) | 591 (40.6%) | 615 (33.9%) | 66 (45.5%) | 1393 (37.0%) |
| Hospital admissions prior<br>year (mean, SD) | 1.51 (2.90) | 1.26 (2.46) | 1.32 (2.46) | 1.31 (2.90) | 1.72 (4.70) | 1.37 (3.70) | 1.65 (3.29) | 1.32 (2.87) |
| Prescriptions prior year<br>(mean, SD) | 3.86 (3.10) | 3.46 (3.10) | 3.40 (2.85) | 3.33 (3.10) | 3.35 (2.93) | 3.20 (2.98) | 3.12 (2.65) | 3.29 (3.14) |

### Readmission with stroke

Over follow-up, 5,621 (8.71%) of people with stroke/SDH were subsequently admitted with stroke/SDH. Readmission with stroke within 6 months occurred in 2.7% to 2.9% of people with ischemic stroke, and 2.3% to 4.6% with ICH (Supplement 1, eFigure 4), varying by age and sex. Those with lobar ICH were more likely to be readmitted than those with deep ICH (aHR 1.71 (1.15-2.54), Figure 1) but there was no difference between those with cortical ischemic stroke compared with deep (aHR 1.14 (0.99-1.31)). Beyond 6 months, there were no significant differences between stroke subtypes.

**Figure 1:**
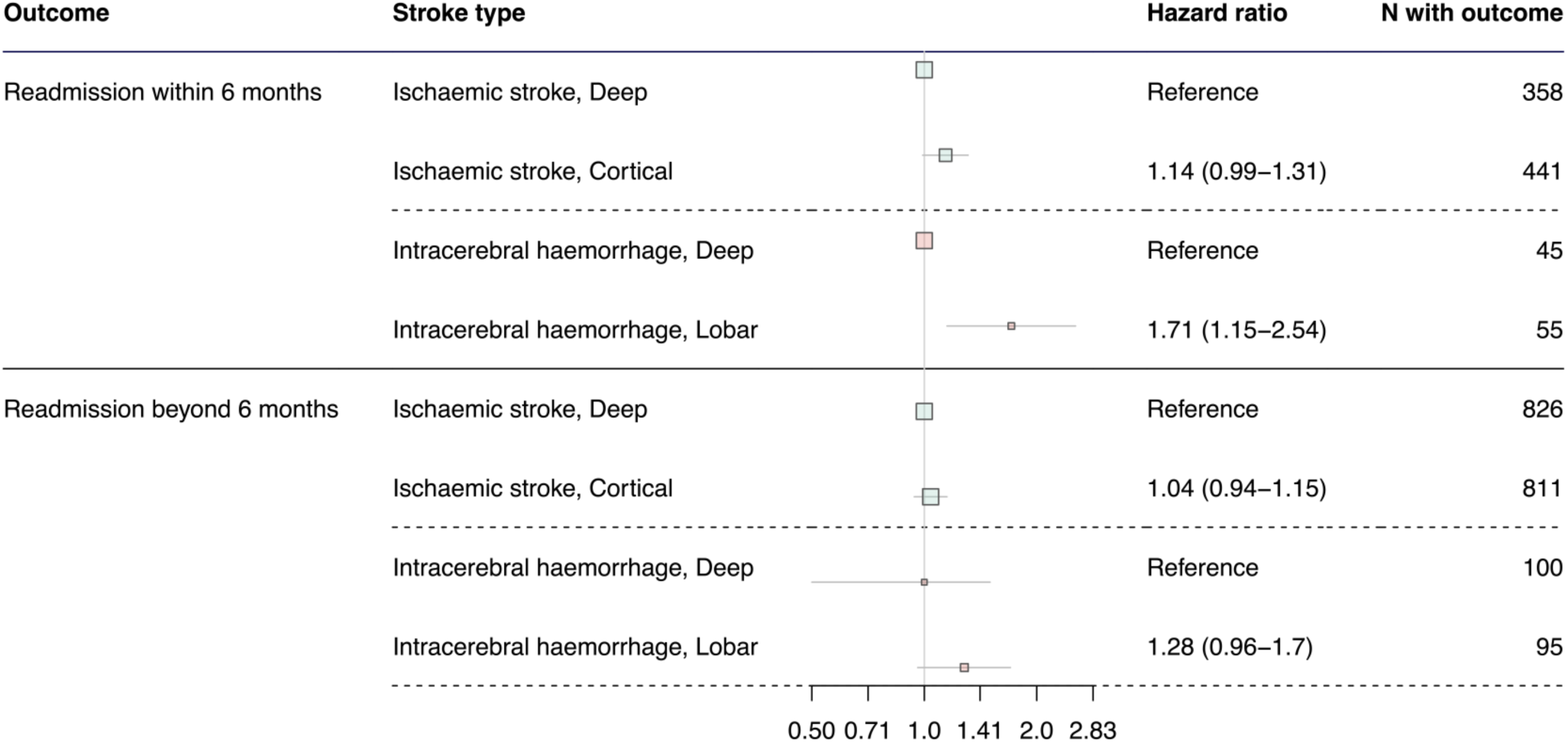
Fully adjusted hazard ratios for readmission with stroke subtypes. Deep ischemic stroke is compared with cortical ischemic stroke; deep intracerebral hemorrhage is compared with lobar intracerebral hemorrhage.

### MI

During follow-up, 2680 of people with stroke/SDH had MI (number rounded to nearest 10 for data suppression purposes). The aHR for MI early after stroke (<6 months) was highest for those with ischemic stroke (aHR 3.35 (2.79-4.03)) and SDH (aHR 3.97 (1.73-9.1)). Among subtypes, the highest aHR was for those with cortical ischemic stroke (aHR 4.6 (3.35-6.31), Figure 2).

**Figure 2:**
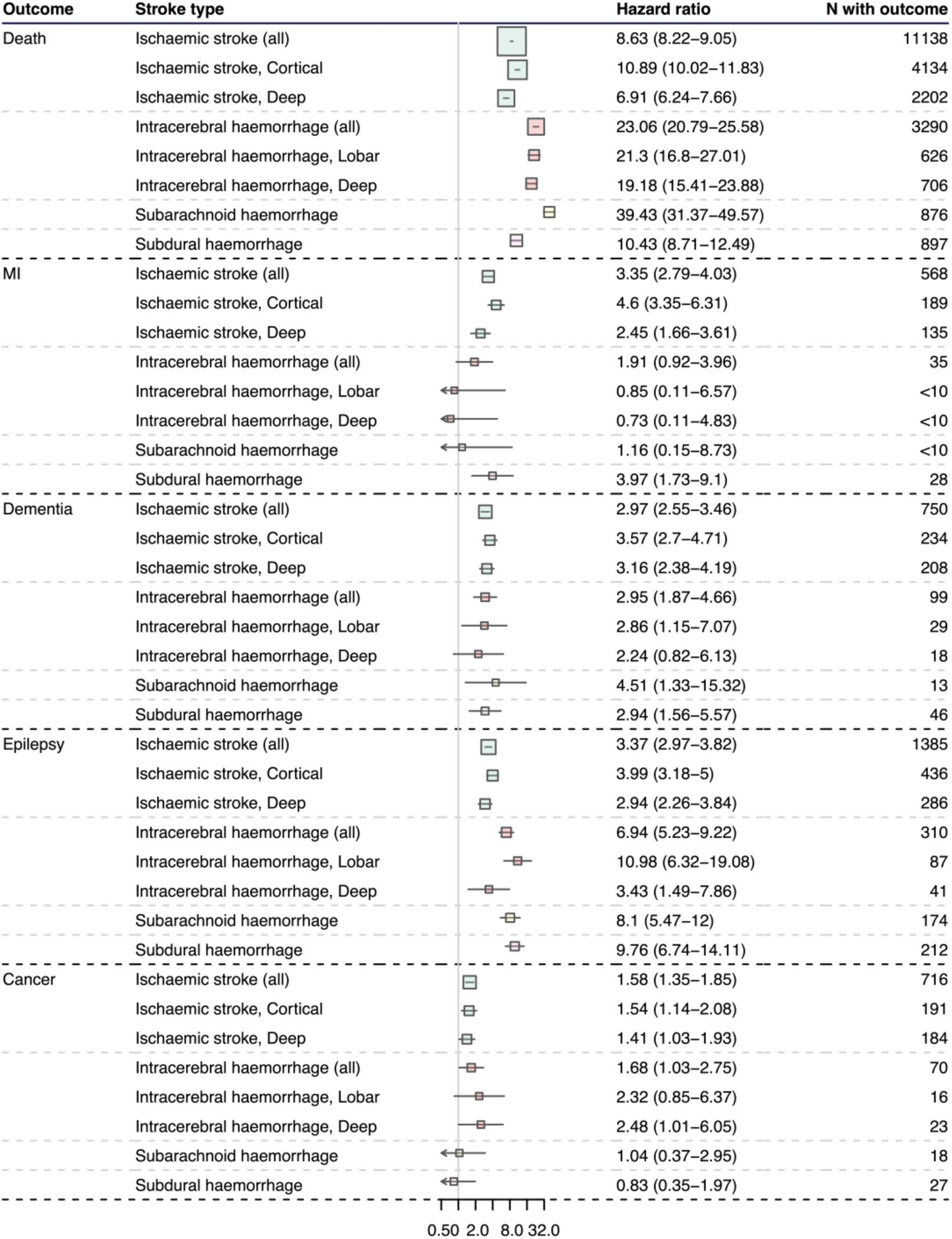
Fully adjusted hazard ratios for death, myocardial infarction, dementia, epilepsy and cancer early after stroke (<6 months), compared to age- and sex-matched controls without stroke. Abbreviations: MI=myocardial infarction, IS=ischemic stroke, ICH=intracerebral hemorrhage, SAH=subarachnoid hemorrhage, SDH=subdural hemorrhage.

### Dementia

5139/59720 (8.6%) of people with stroke, without pre-existing dementia, developed dementia. Early after stroke (<6 months), the aHR for dementia was similar for stroke types and subtypes. Late after stroke (>6 months), compared with matched controls, the aHR for lobar ICH (3.49 (2.3-5.29)) was higher than for deep ICH (2.27 (1.47-3.49)), cortical ischemic stroke (2.16 (1.9-2.46)) and deep ischemic stroke (1.94 (1.71-2.21)) (Figure 3). Accounting for competing risk of death, cumulative incidence of dementia by 5 years in men over 70 years was 17% for lobar ICH, 6.5% for deep ICH, 11% for cortical ischemic stroke, and 10% for deep ischemic stroke (Figure 4).

**Figure 3:**
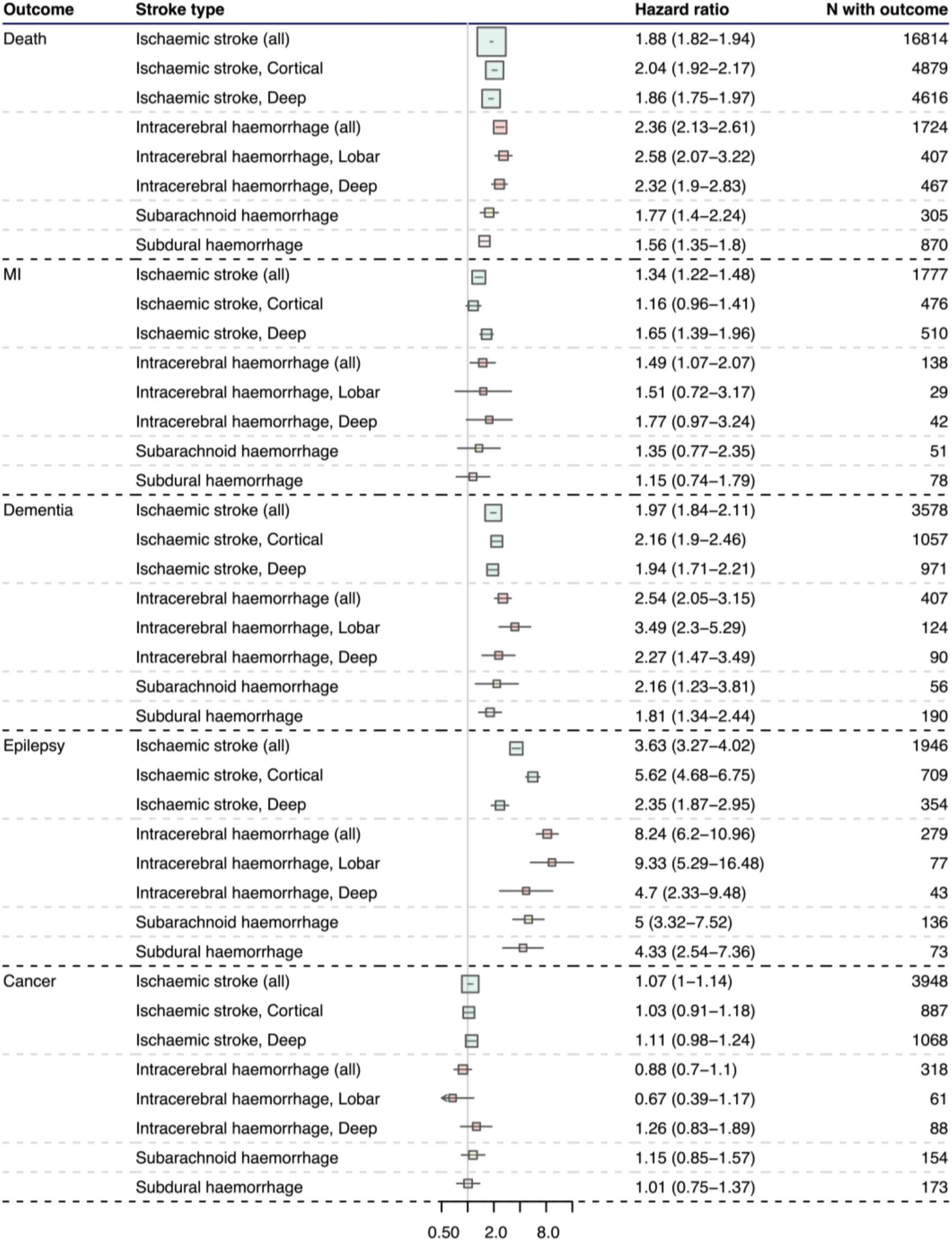
Fully adjusted hazard ratios for death, myocardial infarction, dementia, epilepsy and cancer, late after stroke (beyond 6 months), compared with age- and sex-matched controls without stroke. Abbreviations: MI=myocardial infarction, IS=ischemic stroke, ICH=intracerebral hemorrhage, SAH=subarachnoid hemorrhage, SDH=subdural hemorrhage.

**Figure 4:**
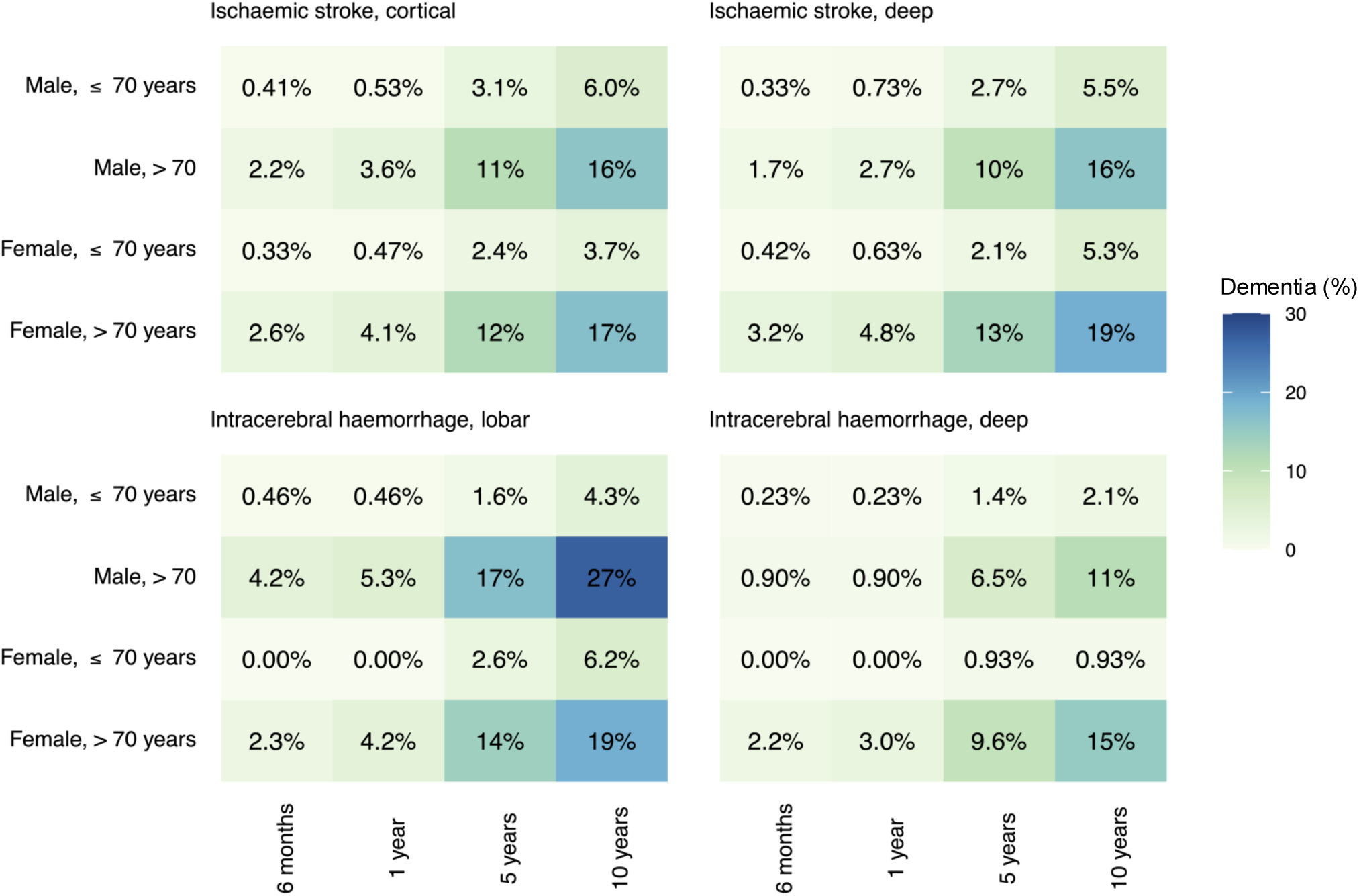
Cumulative incidence rates dementia accounting for competing risk of death (B) for ischemic stroke and intracerebral hemorrhage by location, stratified by age and sex.

### Cancer

During follow-up, 5424/56610 (9.6%) of people with stroke without pre-existing cancer, developed cancer compared with 13.1% of matched controls. Early after stroke (<6 months), there was elevated cancer risk for people with cortical ischemic stroke (aHR 1.54 (1.14-2.08); deep ischemic stroke (aHR 1.41 (1.03-1.93), and deep ICH (aHR 2.48 (1.01-6.05)). The aHR for lobar ICH was consistent with no increase (2.32 (0.85-6.37), Figure 2). Beyond 6 months, we detected no difference in cancer incidence between people without stroke and people with any stroke subtypes (Figure 3).

### Epilepsy

During follow-up, 4515/62548 (7.2%) of people with stroke, without preexisting epilepsy, developed epilepsy. Cumulative incidence, accounting for competing risk of death, was higher for those with cortical and lobar strokes (eFigure 4). Early after stroke the aHR for epilepsy was higher for cortical than deep ischemic stroke (3.99 (3.18-5) vs 2.94 (2.26-3.84)), and for lobar than for deep ICH (10.98 (6.82-19.08) vs 3.42 (1.49-7.86), Figure 2). Late after stroke (>6 months), the same pattern was seen among subtypes, although the aHRs were generally greater (Figure 3).

### Death

During follow-up, 36,914 (55.9%) of people with stroke died (Supplement 1, eFigure 5). Almost half (45.8%) of those with ICH died in the first 6 months, compared to 21.7% with ischemic stroke. Both short- and long-term survival declined with increasing age (Figure 5).

**Figure 5:**
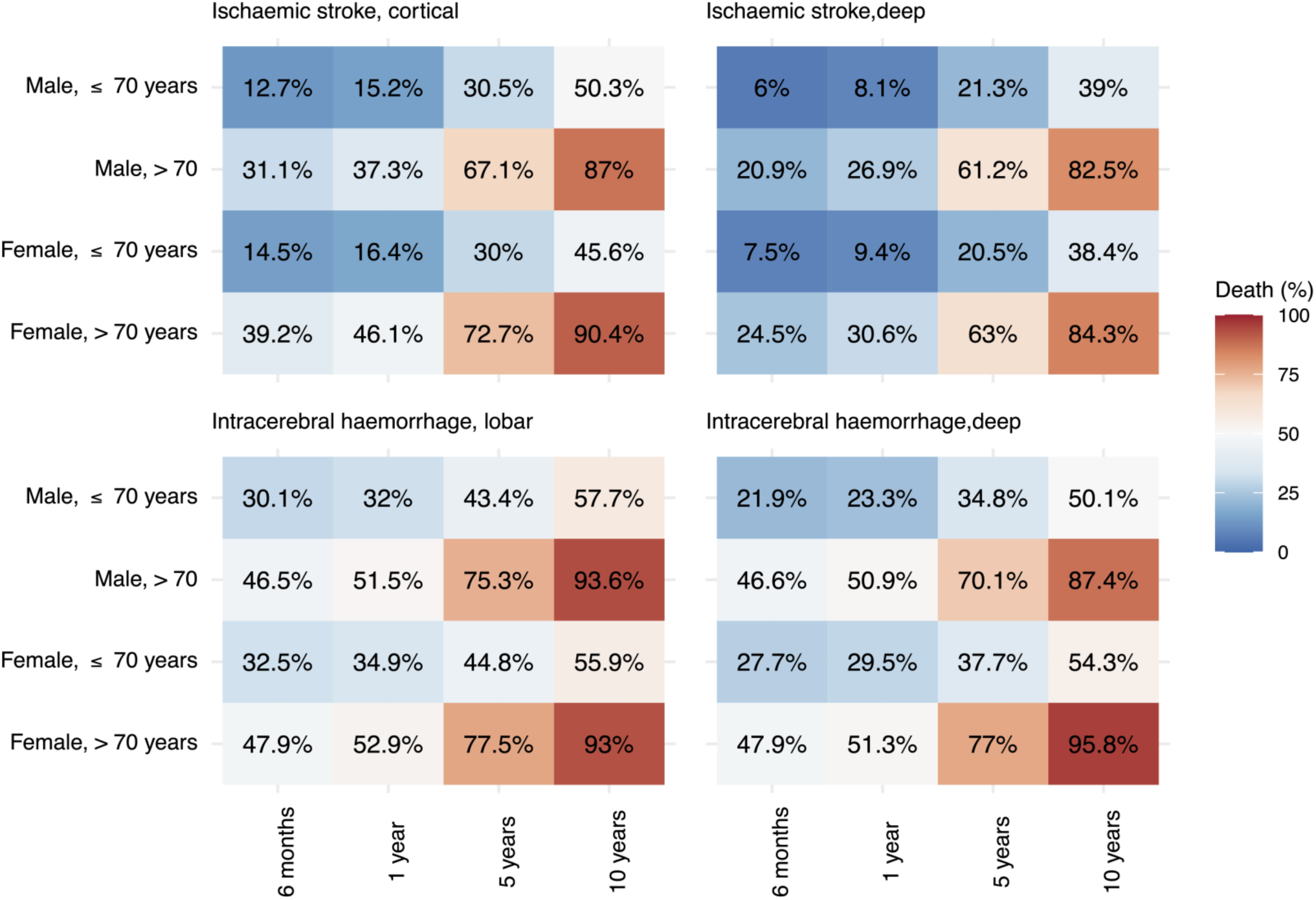
Cumulative incidence rates of death for ischemic stroke and intracerebral hemorrhage by location, stratified by age and sex.

Early after stroke (<6 months), compared with matched controls, the aHR for death was higher for ICH than ischemic stroke (23.06 (20-79-25.58) vs 8.63 (8.22-9.05)), and the aHR for death was higher for cortical than deep ischemic stroke (10.9 (10.02-11.83) vs 6.91 (6.24-7,66)). Lobar and deep ICH had similar aHR (21.3 (16.8-27.0) vs 19.2 (15.4-23.9)), Figure 2). Beyond 6 months, cortical ischemic stroke continued to have higher aHR for death than deep ischemic stroke (2.04 (1.92-2.17) vs 1.86 (1.75-1.97)) (Figure 3).

The most common cause of death among people with ischemic stroke was unspecified stroke; for ICH, SAH and SDH, it was the recorded stroke type. Among matched controls, the most common cause of death was unspecified dementia (Supplement 1, eTable 4). 11,774 people with stroke/SDH died had stroke/SDH as underlying cause of death, of whom 7,609 (65.6%) died in the first month after stroke.

## Discussion

In linked healthcare systems data from NHS Scotland, we studied stroke subtypes at unprecedented scale with bespoke NLP of radiology reports. We reduced the number of people with unknown stroke types from 17,472 to 2,618. We confirmed the high rate of early mortality for ICH compared to ischemic stroke; higher long-term risk of term dementia after lobar ICH than for other stroke subtypes; higher short- and long-term risk of epilepsy for cortical and lobar subtypes than for deep subtypes; and higher short-term risk of MI for cortical ischemic stroke than for other subtypes. These findings validate our NLP method for use in research and monitoring of care pathways.

We defined readmission with stroke only when a stroke code was in the primary position, as a proxy for stroke recurrence. We did not include death records of stroke or SDH without a corresponding hospital admission, as these likely represent death from the original stroke. Readmission was increased for those with lobar ICH compared to deep ICH. Our results are consistent with previous findings: in 674 people from two pooled British cohorts, recurrence rates were higher in those with lobar ICH (RR 2.8 (1.5-5.5)).^1^

We found an association between cortical ischemic stroke and early MI (aHR 4.6 (3.35-6.31). A previous systematic review found no difference in cardiac death in those with lacunar and non-lacunar stroke, but based on relatively few events.^16^ Cortical ischemic stroke and MI may share pathological origins of large artery artherosclerosis.

People with stroke did not have long-term increased risk of cancer in this study and only a modest early elevated risk. In the Canadian Longitudinal Health study, there was similarly increased risk of cancer in the first year after stroke but not thereafter, compared with age-matched controls without stroke;^17^ and the Women’s Health Initiative found stroke was associated with a lower risk of subsequent cancer in post-menopausal women.^18^

Long-term dementia risk was increased after stroke, particularly lobar ICH (aHR 3.48 (2.3-5.29)). The same pattern was found in single center cohorts of ICH in the US and France.^19,20^ Our detection rates of dementia are lower than cohort studies, presumably because most dementia is diagnosed in outpatient settings (10.5% of a population-based cohort of people with stroke in England developed dementia after one year, compared to 2.5% of our cohort).^21^ Epilepsy risk was elevated for the first 6 months after stroke and remained elevated beyond 6 months, particularly for cortical and lobar locations, consistent with previous data.^22–24^

Strengths of our study were large size, representativeness of clinical practice, and inclusion of a whole-country health-system including academic and non-academic centers. We used an NLP algorithm with good positive predictive value for ischemic and hemorrhagic stroke, and stroke location, allowing us to reclassify most people with non-specific stroke codes, and to create large cohorts of stroke subtypes by location.^13^ Our innovative NLP approach can, with appropriate algorithms, be applied to other conditions.^25^

Limitations were that we could not identify subtypes for around half of cases of ICH and ischemic stroke, although the aHR for those with no location were similar to those with identified locations (Supplement 1, eFigure 6). With direct access to images, an algorithm could identify stroke subtypes without radiology reports, potentially reducing the number with unknown location subtypes.^26^ Our NLP method was rules-based, whereas most modern systems employ large language models. We did not use these systems as the governance requirements in secure data environments are onerous, and they may have similar performance to smaller, expert developed models.^27^ NLP identified atrophy and WMH in scan reports, but these are not adjusted for as they are more likely to be recorded in a person with stroke (who therefore had at least one head scan), compared to the control population who may not have had a scan.

We have previously validated EdiE-R’s categorization of stroke subtypes, but did not validate our process of comparing ICD-10 codes and NLP tags for this study. However, baseline characteristics aligned with our understanding of underlying pathology of subtypes.^28,29^ Those with cortical ischemic stroke had highest rates of atrial fibrillation (18.9% vs 12.1% for deep ischemic stroke), and those with deep ischemic stroke had higher rates of atrophy and WMH than those with cortical. An alternative study design would use time-varying covariates; however, we chose a matched cohort study as for a cohort of our size with variables updated daily, time-varying covariates are computationally unfeasible. We chose cause-specific models rather than competing risk models as they allow estimation of effects of covariates on the rate of occurrence of the outcome in those subjects who are currently event free, and are suited to addressing etiological questions, although we accounted for competing risk when estimating cumulative incidence.^30^

We used real-world, nationwide, healthcare-data enhanced with NLP of imaging reports to produce estimates for risks of outcomes directly relevant to people with stroke subtypes. Our approach is one example of how NLP and other artificial intelligence technologies allow analysis of large volumes of data without requiring expert input for each individual case, with enormous possibilities for audit and epidemiology. Artificial intelligence tools already assist in imaging analysis and clinical decision making in stroke medicine.^31^ Beyond routine clinical care, automated imaging analysis could identify phenotypes in large-scale national cohort studies, such as UK Biobank and Our Future Health,^32,33^ and identify participants for trials of interventions for stroke and other conditions.

## Supporting information

Supplement 2 - code lists

## Acknowledgements

For the purpose of open access, the author(s) has applied a Creative Commons attribution (CC BY) licence to any Author Accepted Manuscript version arising.

## Author Contributions

Alice Hosking had full access to all the data in the study and takes responsibility for the integrity of the data and the accuracy of the data analysis.

WNW and AH conceived the study. AH conducted the statistical analysis. BA, CG, and RT implemented the natural language processing algorithm and extracted phenotype data. WNW, MI, and LS provided methodological and statistical advice. AH and WNW drafted the manuscript. All authors reviewed the manuscript and provided feedback.

## Sources of funding and support

AH is funded by a Medical Research Council (UK)/The Stroke Association fellowship (MR/Z504051/1). This project was funded by the Chief Scientist’s Office (CSO-SCAF/17/01), the Medical Research Council (G0902303/1), the Alzheimer’s Society (486). WNW is supported by CSO and Health Data Research UK. MHI is supported by the Wellcome Trust (220857/Z/20/Z; 226770/Z/22/Z, 104036/Z/14/Z; 216767/Z/19/Z) and by a Research Data Scotland Accelerator Award (RAS-24-2). MM is funded by Health Data Research UK. BA is supported by the Turing Fellowship and Turing project (EP/N510129/1) from The Alan Turing Institute, by Legal and General PLC as part of the Advanced Care Research Centre, and by the National Institute for Health Research (NIHR202639). JMW is part funded by the UK DRI which is funded by UK MRC, Als Soc and ARUK; JMW is also part funded by NIHR.

## Conflict of Interest Disclosures

GM has received consultancy fees from Canon Medical Research, Europe, Ltd. The other authors declare no other conflicts of interest

## Notes

### Summary of Updates

Added Supplementary Material not included with original upload.

## References

1. Li L, Poon MTC, Samarasekera NE, et al. Risks of recurrent stroke and all serious vascular events after spontaneous intracerebral haemorrhage: pooled analyses of two population-based studies. Lancet Neurol. 2021;20(6):437–447. doi:10.1016/S1474-4422(21)00075-2

2. Verburgt E, Fellah L, Ekker MS, et al. Risk of Poststroke Epilepsy Among Young Adults With Ischemic Stroke or Intracerebral Hemorrhage. JAMA Neurol. 2025;82(6):597–604. doi:10.1001/JAMANEUROL.2025.0465

3. Wang Y, Jing J, Meng X, et al. The Third China National Stroke Registry (CNSR-III) for patients with acute ischaemic stroke or transient ischaemic attack: design, rationale and baseline patient characteristics. Stroke Vasc Neurol. 2019;4(3):158–164. doi:10.1136/SVN-2019-000242

4. Skajaa N, Adelborg K, Horváth-Puhó E, et al. Nationwide Trends in Incidence and Mortality of Stroke Among Younger and Older Adults in Denmark. Neurology. 2021;96(13):E1711–E1723. doi:10.1212/WNL.0000000000011636/ASSET/C2ACC646-606D-4FEC-95B2-62DA91FD81BD/ASSETS/GRAPHIC/6TTU1.JPEG

5. Skajaa N, Adelborg K, Horváth-Puhó E, et al. Risks of Stroke Recurrence and Mortality After First and Recurrent Strokes in Denmark: A Nationwide Registry Study. Neurology. 2022;98(4):E329–E342. doi:10.1212/WNL.0000000000013118

6. Bray BD, Paley L, Hoffman A, et al. Socioeconomic disparities in first stroke incidence, quality of care, and survival: a nationwide registry-based cohort study of 44 million adults in England. Lancet Public Health. 2018;3(4):e185–e193. doi:10.1016/S2468-2667(18)30030-6

7. Jarman JW, Hunter TD, Hussain W, March JL, Wong T, Markides V. Stroke rates before and after ablation of atrial fibrillation and in propensity-matched controls in the UK. Pragmat Obs Res. 2017;8:107–118. doi:10.2147/POR.S134781

8. Woodfield R, Grant I, Group UBSO, Group UBFU and OW, Sudlow CLM. Accuracy of Electronic Health Record Data for Identifying Stroke Cases in Large-Scale Epidemiological Studies: A Systematic Review from the UK Biobank Stroke Outcomes Group. PLoS One. 2015;10(10):e0140533. doi:10.1371/JOURNAL.PONE.0140533

9. Roark C, Wilson MP, Kubes S, Mayer D, Wiley LK. Assessing the utility and accuracy of ICD10-CM non-traumatic subarachnoid hemorrhage codes for intracranial aneurysm research. Learn Health Syst. 2021;5(4):e10257. doi:10.1002/LRH2.10257

10. Rannikmäe K, Ngoh K, Bush K, et al. Accuracy of identifying incident stroke cases from linked health care data in UK Biobank. Neurology. 2020;95(6):E697–E707. doi:10.1212/WNL.0000000000009924

11. Wheater E, Mair G, Sudlow C, Alex B, Grover C, Whiteley W. A validated natural language processing algorithm for brain imaging phenotypes from radiology reports in UK electronic health records. BMC Med Inform Decis Mak. 2019;19(1):1–11. doi:10.1186/S12911-019-0908-7/TABLES/5

12. Alex B, Grover C, Tobin R, Sudlow C, Mair G, Whiteley W. Text mining brain imaging reports. J Biomed Semantics. 2019;10(1):1–11. doi:10.1186/S13326-019-0211-7/TABLES/8

13. Casey A, Davidson E, Grover C, et al. Understanding the performance and reliability of NLP tools: a comparison of four NLP tools predicting stroke phenotypes in radiology reports. Frontiers in Digital Health, 5, Article 1184919 (2023). 2023;5. doi:10.3389/FDGTH.2023.1184919

14. Baxter R, Nind T, Sutherland J, et al. The Scottish Medical Imaging Archive: 57.3 Million Radiology Studies Linked to Their Medical Records. Radiol Artif Intell. 2023;6(1):e220266. doi:10.1148/RYAI.220266

15. Migration flows - National Records of Scotland (NRS). Accessed December 19, 2024. https://www.nrscotland.gov.uk/publications/migration-flows/

16. Jackson C, Sudlow C. Comparing risks of death and recurrent vascular events between lacunar and non-lacunar infarction. Brain. 2005;128(Pt 11):2507. doi:10.1093/BRAIN/AWH636

17. Rioux B, Gioia LC, Keezer MR. Risk of Cancer Following an Ischemic Stroke in the Canadian Longitudinal Study on Aging. Can J Neurol Sci. 2022;49(2):225–230. doi:10.1017/CJN.2021.55

18. Sealy-Jefferson S, Cote ML, Chlebowski RT, Rexrode KM, Simon MS. Post-Stroke Cancer Risk among Postmenopausal Women: The Women’s Health Initiative. Womens Health Issues. 2018;28(1):29–34. doi:10.1016/J.WHI.2017.10.012

19. Biffi A, Bailey D, Anderson CD, et al. Risk Factors Associated With Early vs Delayed Dementia After Intracerebral Hemorrhage. JAMA Neurol. 2016;73(8):969–976. doi:10.1001/JAMANEUROL.2016.0955

20. Moulin S, Labreuche J, Bombois S, et al. Dementia risk after spontaneous intracerebral haemorrhage: a prospective cohort study. Lancet Neurol. 2016;15(8):820–829. doi:10.1016/S1474-4422(16)00130-7

21. Pendlebury ST, Rothwell PM. Incidence and prevalence of dementia associated with transient ischaemic attack and stroke: analysis of the population-based Oxford Vascular Study. Lancet Neurol. 2019;18(3):248. doi:10.1016/S1474-4422(18)30442-3

22. Biffi A, Rattani A, Anderson CD, et al. Delayed seizures after intracerebral haemorrhage. Brain. 2016;139(Pt 10):2694–2705. doi:10.1093/BRAIN/AWW199

23. Galovic M, Döhler N, Erdélyi-Canavese B, et al. Prediction of late seizures after ischaemic stroke with a novel prognostic model (the SeLECT score): a multivariable prediction model development and validation study. Lancet Neurol. 2018;17(2):143. doi:10.1016/S1474-4422(17)30404-0

24. Graham NSN, Crichton S, Koutroumanidis M, Wolfe CDA, Rudd AG. Incidence and associations of poststroke epilepsy the prospective South London stroke register. Stroke. 2013;44(3):605–611. doi:10.1161/STROKEAHA.111.000220/SUPPL_FILE/STR202675_SUPPLEMENT.PDF

25. Iveson MH, Mukerjee M, Davidson EM, et al. Clinically reported covert cerebrovascular disease and risk of neurological disease: a whole-population cohort of 395,273 people using natural language processing. medRxiv. Published online June 23, 2025:2025.06.12.25329472. doi:10.1101/2025.06.12.25329472

26. Camilleri MPJ, Gouzou D, Al-Wasity S, et al. A large dataset of brain imaging linked to health systems data: the curation and access to a whole system national cohort from NHS Scotland. medRxiv. Published online October 24, 2025:2025.10.21.25338469. doi:10.1101/2025.10.21.25338469

27. Peng Y, Wang X, Lu L, Bagheri M, Summers R, Lu Z. NegBio: a high-performance tool for negation and uncertainty detection in radiology reports. AMIA Jt Summits Transl Sci Proc. 2018;2017(2):188–196. Accessed January 20, 2026. http://www.ncbi.nlm.nih.gov/pubmed/29888070

28. Dupré N, Drieu A, Joutel A. Pathophysiology of cerebral small vessel disease: a journey through recent discoveries. J Clin Invest. 2024;134(10). doi:10.1172/JCI172841

29. Kamel H, Healey JS. Cardioembolic Stroke. Circ Res. 2017;120(3):514. doi:10.1161/CIRCRESAHA.116.308407

30. Austin PC, Lee DS, Fine JP. Introduction to the Analysis of Survival Data in the Presence of Competing Risks. Circulation. 2016;133(6):601–609. doi:10.1161/CIRCULATIONAHA.115.017719/-/DC1

31. Al-Janabi OM, El Refaei A, Elgazzar T, et al. Current Stroke Solutions Using Artificial Intelligence: A Review of the Literature. Brain Sci. 2024;14(12):1182. doi:10.3390/BRAINSCI14121182/S1

32. Cook MB, Adams N, Adjetey A, et al. Cohort Profile: Our Future Health. Int J Epidemiol. 2025;54(6):dyaf171. doi:10.1093/IJE/DYAF171

33. Sudlow C, Gallacher J, Allen N, et al. UK biobank: an open access resource for identifying the causes of a wide range of complex diseases of middle and old age. PLoS Med. 2015;12(3). doi:10.1371/JOURNAL.PMED.1001779

