## Supplement 1 for "Prognosis of stroke subtypes in whole population health systems data: a matched cohort study"

relating to

eFigure 1. Flow diagram showing selection of stroke cohort

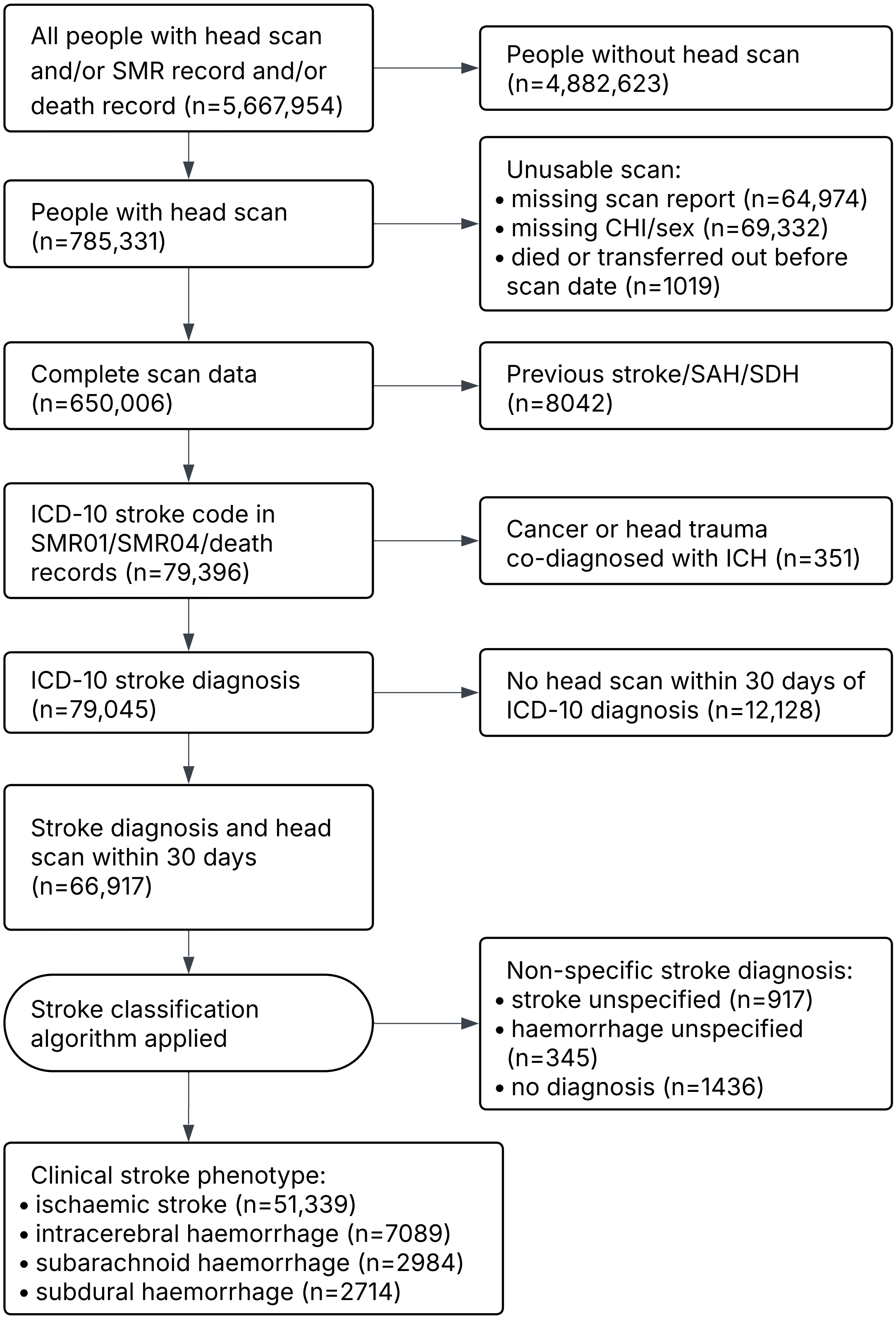

eTable 1. Decision table for reassigning stroke subtypes based on NLP coding

For each ICD-10 code, if the NLP label in the first row existed in a person's scan, they were assigned that final diagnosis. If not, we moved to the next row.

| **Original ICD10 code** | **NLP label** | **Final diagnosis** |
|---|---|---|
| **I63 Cerebral infarction** | Ischaemic stroke, recent | Ischaemic stroke |
| | Haemorrhagic transformation | Ischaemic stroke |
| | ICH, recent | ICH |
| | Ischaemic stroke, old | Ischaemic stroke |
| | Haemorrhage, unspecified | Ischaemic stroke |
| | Ischaemic stroke, unspecified | Ischaemic stroke |
| | Stroke unknown | Ischaemic stroke |
| | ICH, old | ICH |
| | SAH | SAH |
| | SDH | SDH |
| | No label | Ischaemic stroke |
| **I61 Intracerebral haemorrhage** | ICH, recent | ICH |
| | Haemorrhage, unspecified | ICH |
| | Ischaemic stroke, recent | Ischaemic stroke |
| | Haemorrhagic transformation | Ischaemic stroke |
| | ICH, old | ICH |
| | Ischaemic stroke, unspecified | Ischaemic stroke |
| | Ischaemic stroke, old | Ischaemic stroke |
| | SAH | SAH |
| | SDH | SDH |
| | Stroke unknown | ICH |
| | No label | Stroke unspecified |
| **I60 Subarachnoid haemorrhage** | SAH | SAH |
| | Other/no label | No diagnosis |
| **I62.0 Nontraumatic subdural haemorrhage** | SDH | SDH |
| | Other/no label | No diagnosis |

| | | |
|---|---|---|
| **I64 Stroke, not specified as haemorrhage or infarction** | Ischaemic stroke, recent | Ischaemic stroke |
| | Haemorrhagic transformation | Ischaemic stroke |
| | ICH, recent | ICH |
| | SAH | SAH |
| | SDH | SDH |
| | Ischaemic stroke, old | Ischaemic stroke |
| | Haemorrhage, unspecified | Ischaemic stroke |
| | Ischaemic stroke, unspecified | Ischaemic stroke |
| | ICH, old | ICH |
| | Stroke unknown | Stroke unspecified |
| | No label | Ischaemic stroke |
| **I62.9 Intracranial haemorrhage (nontraumatic), unspecified** | ICH, recent | ICH |
| | Haemorrhagic transformation | Ischaemic stroke |
| | SAH | SAH |
| | SDH | SDH |
| | ICH, old | ICH |
| | ICH, unspecified | Haemorrhage unspecified |
| | Other/no label | No diagnosis |

eFigure 2. Matrix showing original ICD-10 codes and final NLP-amended diagnosis

*Final diagnosis* (columns); *ICD-10 code* (rows)

| ICD-10 code | Ischaemic stroke | ICH | SAH | SDH | Haemorrhage unspecified | Stroke unspecified | No diagnosis |
|---|---|---|---|---|---|---|---|
| Ischaemic stroke | 99% | 1% | 0% | 0% | 0% | 0% | 0% |
| ICH | 5% | 87% | 1% | 1% | 0% | 6% | 0% |
| SAH | 0% | 1% | 71% | 0% | 0% | 0% | 28% |
| SDH | 0% | 0% | 0% | 90% | 0% | 0% | 10% |
| Non–traumatic haemorrhage | 3% | 32% | 9% | 17% | 30% | 0% | 9% |
| Stroke unspecified | 94% | 2% | 0% | 1% | 0% | 3% | 0% |

eTable 2. Baseline characteristics of people with stroke and matched controls, controls separated by stroke type

| | Ischaemic stroke | | Intracerebral haemorrhage | | Subarachnoid haemorrhage | | Subdural haemorrhage | |
|---|---|---|---|---|---|---|---|---|
| | **Case (N=51,339)** | **Control (N=205,356)** | **Case (N=7182)** | **Control (N=28,728)** | **Case (N=2984)** | **Control (N=11,936)** | **Case (N=2714)** | **Control (N=10,856)** |
| **Sex** (male) | 25681 (50.0%) | 102724 (50.0%) | 3414 (47.5%) | 13656 (47.5%) | 999 (33.5%) | 3996 (33.5%) | 1695 (62.5%) | 6780 (62.5%) |
| **Age** (years, mean (SD)) | 74.0 (13.4) | 73.8 (13.3) | 73.6 (13.9) | 73.5 (13.8) | 61.1 (14.8) | 61.0 (14.7) | 76.0 (13.6) | 75.8 (13.5) |
| **Atrophy on scan** | 24675 (48.1%) | 27918 (13.6%) | 2894 (40.3%) | 3906 (13.6%) | 402 (13.5%) | 713 (6.0%) | 1260 (46.4%) | 1635 (15.1%) |
| **SVD on scan** | 30557 (59.5%) | 27864 (13.6%) | 3735 (52.0%) | 3920 (13.6%) | 746 (25.0%) | 754 (6.3%) | 1184 (43.6%) | 1596 (14.7%) |
| **Diabetes** | 6636 (12.9%) | 12317 (6.0%) | 691 (9.6%) | 1731 (6.0%) | 105 (3.5%) | 440 (3.7%) | 373 (13.7%) | 700 (6.4%) |
| **Hypertension** | 34168 (66.6%) | 110246 (53.7%) | 4438 (61.8%) | 15451 (53.8%) | 1040 (34.9%) | 4035 (33.8%) | 1841 (67.8%) | 6175 (56.9%) |
| **Atrial fibrillation** | 7444 (14.5%) | 12870 (6.3%) | 875 (12.2%) | 1798 (6.3%) | 115 (3.9%) | 288 (2.4%) | 538 (19.8%) | 813 (7.5%) |
| **Statins** | 23896 (46.5%) | 77859 (37.9%) | 3130 (43.6%) | 10824 (37.7%) | 744 (24.9%) | 2798 (23.4%) | 1327 (48.9%) | 4374 (40.3%) |
| **Anticoagulants** | 5161 (10.1%) | 13650 (6.6%) | 1116 (15.5%) | 1929 (6.7%) | 154 (5.2%) | 399 (3.3%) | 646 (23.8%) | 840 (7.7%) |
| **Antiplatelets** | 21617 (42.1%) | 57864 (28.2%) | 2665 (37.1%) | 8011 (27.9%) | 502 (16.8%) | 1701 (14.3%) | 1054 (38.8%) | 3457 (31.8%) |
| **Hospital admissions in last year (n**, mean (SD)**)** | 1.35 (2.79) | 0.585 (1.71) | 1.42 (3.53) | 0.558 (1.62) | 0.673 (1.92) | 0.394 (1.53) | 2.56 (4.13) | 0.672 (2.40) |
| **Prescriptions in last year (n**, mean (SD)**)** | 3.51 (3.11) | 3.32 (3.25) | 3.28 (3.05) | 3.29 (3.25) | 1.68 (2.48) | 2.36 (2.96) | 3.89 (3.14) | 3.49 (3.27) |

Baseline characteristics of people with stroke and matched controls, controls separated by stroke type, continued...

| | **Stroke unspecified** | | **Haemorrhage unspecified** | | **No diagnosis** | |
|---|---|---|---|---|---|---|
| | **Case (N=917)** | **Control (N=3668)** | **Case (N=345)** | **Control (N=1380)** | **Case (N=1436)** | **Control (N=5744)** |
| **Sex** (male) | 445 (48.5%) | 1780 (48.5%) | 170 (49.3%) | 680 (49.3%) | 641 (44.6%) | 2564 (44.6%) |
| **Age** (years, mean (SD)**)** | 71.6 (14.5) | 71.4 (14.5) | 75.0 (13.3) | 74.8 (13.2) | 62.2 (16.0) | 62.2 (16.0) |
| **Atrophy on scan** | 400 (43.6%) | 393 (10.7%) | 118 (34.2%) | 173 (12.5%) | 344 (24.0%) | 444 (7.7%) |
| **SVD on scan** | 466 (50.8%) | 372 (10.1%) | 162 (47.0%) | 185 (13.4%) | 465 (32.4%) | 456 (7.9%) |
| **Diabetes** | 126 (13.7%) | 227 (6.2%) | 35 (10.1%) | 80 (5.8%) | 100 (7.0%) | 226 (3.9%) |
| **Hypertension** | 575 (62.7%) | 1833 (50.0%) | 206 (59.7%) | 771 (55.9%) | 584 (40.7%) | 2082 (36.2%) |
| **Atrial fibrillation** | 116 (12.7%) | 218 (5.9%) | 42 (12.2%) | 107 (7.8%) | 83 (5.8%) | 194 (3.4%) |
| **Statins** | 424 (46.2%) | 1273 (34.7%) | 162 (47.0%) | 534 (38.7%) | 409 (28.5%) | 1487 (25.9%) |
| **Anticoagulants** | 99 (10.8%) | 220 (6.0%) | 72 (20.9%) | 100 (7.2%) | 107 (7.5%) | 235 (4.1%) |
| **Antiplatelets** | 353 (38.5%) | 907 (24.7%) | 131 (38.0%) | 388 (28.1%) | 332 (23.1%) | 977 (17.0%) |
| **Hospital admissions in last year** (n, mean (SD)) | 1.51 (2.99) | 0.520 (1.50) | 1.49 (2.45) | 0.603 (1.61) | 1.16 (2.24) | 0.427 (1.37) |
| **Prescriptions in last year (**n, mean (SD)) | 3.35 (3.21) | 3.13 (3.32) | 3.39 (2.98) | 3.41 (3.27) | 2.08 (2.78) | 2.51 (3.10) |

eTable 3. Baseline characteristics of people with stroke when defined only by ICD-10 code

| | Ischaemic stroke | | Intracerebral haemorrhage | | Subdural haemorrhage | | Subrachnoid haemorrhage | |
|---|---|---|---|---|---|---|---|---|
| | **Case (N=34739)** | **Control (N=138956)** | **Case (N=7075)** | **Control (N=28300)** | **Case (N=2606)** | **Control (N=10424)** | **Case (N=3883)** | **Control (N=15532)** |
| **Sex** (male) | 17697 (50.9%) | 70788 (50.9%) | 3373 (47.7%) | 13492 (47.7%) | 1620 (62.2%) | 6480 (62.2%) | 1376 (35.4%) | 5504 (35.4%) |
| **Age** (years, mean (SD)**)** | 73.5 (13.5) | 73.3 (13.5) | 73.4 (14.1) | 73.2 (14.0) | 76.0 (13.6) | 75.8 (13.5) | 59.8 (14.6) | 59.8 (14.6) |
| **Atrophy on scan** | 16108 (46.4%) | 18251 (13.1%) | 2871 (40.6%) | 3758 (13.3%) | 1224 (47.0%) | 1585 (15.2%) | 513 (13.2%) | 846 (5.4%) |
| **SVD on scan** | 20273 (58.4%) | 18203 (13.1%) | 3665 (51.8%) | 3752 (13.3%) | 1160 (44.5%) | 1545 (14.8%) | 955 (24.6%) | 897 (5.8%) |
| **Diabetes** | 4373 (12.6%) | 8397 (6.0%) | 698 (9.9%) | 1692 (6.0%) | 367 (14.1%) | 688 (6.6%) | 141 (3.6%) | 543 (3.5%) |
| **Hypertension** | 22880 (65.9%) | 73836 (53.1%) | 4366 (61.7%) | 15123 (53.4%) | 1770 (67.9%) | 5938 (57.0%) | 1307 (33.7%) | 4981 (32.1%) |
| **Atrial fibrillation** | 4883 (14.1%) | 8479 (6.1%) | 868 (12.3%) | 1764 (6.2%) | 521 (20.0%) | 799 (7.7%) | 121 (3.1%) | 332 (2.1%) |
| **Statins** | 15950 (45.9%) | 52306 (37.6%) | 3102 (43.8%) | 10624 (37.5%) | 1265 (48.5%) | 4183 (40.1%) | 916 (23.6%) | 3473 (22.4%) |
| **Anticoagulants** | 3399 (9.8%) | 9145 (6.6%) | 1116 (15.8%) | 1899 (6.7%) | 634 (24.3%) | 824 (7.9%) | 166 (4.3%) | 478 (3.1%) |
| **Antiplatelets** | 14190 (40.8%) | 38330 (27.6%) | 2625 (37.1%) | 7841 (27.7%) | 993 (38.1%) | 3289 (31.6%) | 647 (16.7%) | 2090 (13.5%) |
| **Hospital admissions in last year** (n, mean (SD)) | 1.28 (2.74) | 0.578 (1.72) | 1.45 (3.60) | 0.551 (1.62) | 2.51 (3.99) | 0.677 (2.43) | 0.681 (1.86) | 0.373 (1.43) |
| **Prescriptions in last year (**n, mean (SD)) | 3.43 (3.09) | 3.28 (3.24) | 3.31 (3.08) | 3.28 (3.25) | 3.87 (3.12) | 3.49 (3.26) | 1.60 (2.45) | 2.28 (2.93) |

Baseline characteristics of people with stroke when defined only by ICD-10 code, continued...

| | **Non-traumatic haemorrhage unspecified** | | **Stroke unspecified** | |
|---|---|---|---|---|
| | **Case (N=1142)** | **Control (N=4568)** | **Case (N=17472)** | **Control (N=69888)** |
| **Sex** (male) | 563 (49.3%) | 2252 (49.3%) | 8416 (48.2%) | 33664 (48.2%) |
| **Age** (years, mean (SD)**)** | 74.4 (14.0) | 74.2 (13.9) | 75.0 (13.0) | 74.8 (12.9) |
| **Atrophy on scan** | 427 (37.4%) | 663 (14.5%) | 8950 (51.2%) | 10079 (14.4%) |
| **SVD on scan** | 504 (44.1%) | 669 (14.6%) | 10758 (61.6%) | 10081 (14.4%) |
| **Diabetes** | 106 (9.3%) | 259 (5.7%) | 2381 (13.6%) | 4142 (5.9%) |
| **Hypertension** | 704 (61.6%) | 2451 (53.7%) | 11825 (67.7%) | 38264 (54.8%) |
| **Atrial fibrillation** | 157 (13.7%) | 295 (6.5%) | 2663 (15.2%) | 4619 (6.6%) |
| **Statins** | 521 (45.6%) | 1720 (37.7%) | 8338 (47.7%) | 26843 (38.4%) |
| **Anticoagulants** | 215 (18.8%) | 295 (6.5%) | 1825 (10.4%) | 4732 (6.8%) |
| **Antiplatelets** | 444 (38.9%) | 1293 (28.3%) | 7755 (44.4%) | 20462 (29.3%) |
| **Hospital admissions in last year** (n, mean (SD)) | 1.83 (3.23) | 0.596 (1.70) | 1.48 (2.88) | 0.601 (1.67) |
| **Prescriptions in last year (**n, mean (SD)) | 3.42 (3.00) | 3.35 (3.26) | 3.65 (3.14) | 3.40 (3.26) |

### eFigure 3. Heatmaps of cumulative incidence of outcomes for stroke types, stratified by age and sex

#### Cumulative incidence of death

Ischaemic stroke

| | 6 months | 1 year | 5 years | 10 years |
|---|---|---|---|---|
| Male, ≤ 70 years | 8.2% | 10.2% | 22.4% | 38.9% |
| Male, > 70 | 25.4% | 31.2% | 61.6% | 82.9% |
| Female, ≤ 70 years | 9.6% | 11.3% | 21.9% | 36% |
| Female, > 70 years | 30.7% | 36.9% | 65.7% | 85.7% |

Intracerebral haemorrhage

| | 6 months | 1 year | 5 years | 10 years |
|---|---|---|---|---|
| Male, ≤ 70 years | 29% | 30.7% | 41.2% | 51.6% |
| Male, > 70 | 51.7% | 56.1% | 75.8% | 90.3% |
| Female, ≤ 70 years | 35.6% | 37.2% | 45.5% | 55.7% |
| Female, > 70 years | 53.7% | 58% | 79.6% | 94.7% |

Subarachnoid haemorrhage

| | 6 months | 1 year | 5 years | 10 years |
|---|---|---|---|---|
| Male, ≤ 70 years | 19.1% | 19.8% | 22.8% | 30.6% |
| Male, > 70 | 54.9% | 57.9% | 73.5% | 89.2% |
| Female, ≤ 70 years | 19.9% | 20.1% | 23.6% | 30.6% |
| Female, > 70 years | 52.2% | 54.9% | 67% | 79.8% |

Subdural haemorrhage

| | 6 months | 1 year | 5 years | 10 years |
|---|---|---|---|---|
| Male, ≤ 70 years | 17.6% | 19.5% | 33.8% | 45.4% |
| Male, > 70 | 35.9% | 40% | 67.4% | 83.2% |
| Female, ≤ 70 years | 19.6% | 24.6% | 38.2% | 52.6% |
| Female, > 70 years | 42.7% | 48.2% | 74.8% | 90.1% |

#### Cumulative incidence of readmission with stroke, accounting for competing risk of death

Iscahemic stroke

| | 6 months | 1 year | 5 years | 10 years |
|---|---|---|---|---|
| Male, ≤ 70 years | 2.9% | 3.8% | 8.3% | 12% |
| Male, > 70 | 2.9% | 4.0% | 8.5% | 11% |
| Female, ≤ 70 years | 2.7% | 3.6% | 7.5% | 11% |
| Female, > 70 years | 2.8% | 3.8% | 7.9% | 9.7% |

Intracerebral haemorrhage

| | 6 months | 1 year | 5 years | 10 years |
|---|---|---|---|---|
| Male, ≤ 70 years | 4.6% | 5.1% | 8.3% | 12% |
| Male, > 70 | 2.6% | 3.3% | 7.5% | 8.9% |
| Female, ≤ 70 years | 3.5% | 4.1% | 8.0% | 11% |
| Female, > 70 years | 2.3% | 3.7% | 7.0% | 8.1% |

Subarachnoid haemorrhage

| | 6 months | 1 year | 5 years | 10 years |
|---|---|---|---|---|
| Male, ≤ 70 years | 7.0% | 7.4% | 8.8% | 10% |
| Male, > 70 | 4.9% | 6.0% | 9.3% | 13% |
| Female, ≤ 70 years | 6.7% | 7.0% | 8.3% | 10% |
| Female, > 70 years | 6.3% | 7.1% | 10% | 13% |

Subdural haemorrhage

| | 6 months | 1 year | 5 years | 10 years |
|---|---|---|---|---|
| Male, ≤ 70 years | 5.1% | 5.5% | 8.3% | 8.3% |
| Male, > 70 | 5.8% | 6.1% | 9.4% | 10% |
| Female, ≤ 70 years | 5.4% | 5.8% | 7.5% | 11% |
| Female, > 70 years | 6.3% | 8.6% | 12% | 13% |

#### Cumulative incidence of cancer, accounting for competing risk of death

Ischaemic stroke

| | 6 months | 1 year | 5 years | 10 years |
|---|---|---|---|---|
| Male, ≤ 70 years | 1.2% | 1.8% | 6.0% | 12% |
| Male, > 70 | 1.7% | 2.8% | 8.9% | 13% |
| Female, ≤ 70 years | 1.2% | 2.0% | 6.3% | 10% |
| Female, > 70 years | 1.2% | 1.9% | 5.6% | 8.2% |

Intracerebral haemorrhage

| | 6 months | 1 year | 5 years | 10 years |
|---|---|---|---|---|
| Male, ≤ 70 years | 1.6% | 2.1% | 4.9% | 8.1% |
| Male, > 70 | 0.97% | 1.8% | 4.7% | 7.0% |
| Female, ≤ 70 years | 0.75% | 0.97% | 3.6% | 7.2% |
| Female, > 70 years | 0.71% | 1.2% | 2.9% | 4.1% |

Subarachnoid haemorrhage

| | 6 months | 1 year | 5 years | 10 years |
|---|---|---|---|---|
| Male, ≤ 70 years | 0.41% | 0.68% | 3.4% | 8.9% |
| Male, > 70 | 1.5% | 2.3% | 6.8% | 11% |
| Female, ≤ 70 years | 0.51% | 0.72% | 3.0% | 6.6% |
| Female, > 70 years | 0.33% | 1.3% | 3.7% | 6.6% |

Subdural haemorrhage

| | 6 months | 1 year | 5 years | 10 years |
|---|---|---|---|---|
| Male, ≤ 70 years | 0.85% | 0.85% | 4.1% | 6.4% |
| Male, > 70 | 1.9% | 2.8% | 8.5% | 11% |
| Female, ≤ 70 years | 1.9% | 2.7% | 5.6% | 8.1% |
| Female, > 70 years | 1.3% | 1.8% | 4.4% | 5.8% |

#### Cumulative incidence of epilepsy, accounting for competing risk of death

Ischaemic stroke

| | 6 months | 1 year | 5 years | 10 years |
|---|---|---|---|---|
| Male, ≤ 70 years | 3.3% | 5.0% | 8.2% | 9.4% |
| Male, > 70 | 2.5% | 3.4% | 5.4% | 6.1% |
| Female, ≤ 70 years | 3.5% | 4.6% | 8.6% | 11% |
| Female, > 70 years | 2.4% | 3.4% | 5.4% | 6.1% |

Intracerebral haemorrhage

| | 6 months | 1 year | 5 years | 10 years |
|---|---|---|---|---|
| Male, ≤ 70 years | 6.5% | 8.0% | 12% | 14% |
| Male, > 70 | 3.3% | 4.5% | 6.9% | 7.5% |
| Female, ≤ 70 years | 7.5% | 10% | 14% | 16% |
| Female, > 70 years | 3.2% | 4.1% | 5.6% | 6.0% |

Subarachnoid haemorrhage

| | 6 months | 1 year | 5 years | 10 years |
|---|---|---|---|---|
| Male, ≤ 70 years | 6.7% | 8.6% | 12% | 15% |
| Male, > 70 | 3.9% | 3.9% | 4.2% | 5.8% |
| Female, ≤ 70 years | 6.6% | 8.4% | 12% | 13% |
| Female, > 70 years | 4.4% | 4.9% | 6.5% | 7.3% |

Subdural haemorrhage

| | 6 months | 1 year | 5 years | 10 years |
|---|---|---|---|---|
| Male, ≤ 70 years | 14% | 14% | 18% | 19% |
| Male, > 70 | 8.3% | 9.1% | 10% | 11% |
| Female, ≤ 70 years | 7.0% | 7.9% | 11% | 17% |
| Female, > 70 years | 5.8% | 6.5% | 8.0% | 8.7% |

#### Cumulative incidence of dementia, accounting for competing risk of death

Ischaemic stroke

| | 6 months | 1 year | 5 years | 10 years |
|---|---|---|---|---|
| Male, ≤ 70 years | 0.26% | 0.45% | 2.2% | 4.6% |
| Male, > 70 | 2.0% | 3.1% | 11% | 16% |
| Female, ≤ 70 years | 0.30% | 0.45% | 1.9% | 4.0% |
| Female, > 70 years | 2.5% | 4.0% | 12% | 18% |

Intracerebral haemorrhage

| | 6 months | 1 year | 5 years | 10 years |
|---|---|---|---|---|
| Male, ≤ 70 years | 0.21% | 0.28% | 1.7% | 3.2% |
| Male, > 70 | 1.9% | 2.6% | 9.9% | 14% |
| Female, ≤ 70 years | 0.11% | 0.22% | 2.1% | 3.4% |
| Female, > 70 years | 2.6% | 3.6% | 10% | 14% |

Subarachnoid haemorrhage

| | 6 months | 1 year | 5 years | 10 years |
|---|---|---|---|---|
| Male, ≤ 70 years | 0.00% | 0.00% | 0.14% | 1.1% |
| Male, > 70 | 2.1% | 3.4% | 8.6% | 9.4% |
| Female, ≤ 70 years | 0.07% | 0.07% | 0.39% | 0.75% |
| Female, > 70 years | 1.2% | 1.6% | 5.6% | 9.8% |

Subdural haemorrhage

| | 6 months | 1 year | 5 years | 10 years |
|---|---|---|---|---|
| Male, ≤ 70 years | 0.00% | 0.43% | 2.6% | 4.1% |
| Male, > 70 | 2.8% | 3.9% | 11% | 14% |
| Female, ≤ 70 years | 0.39% | 0.79% | 1.2% | 3.4% |
| Female, > 70 years | 2.4% | 4.9% | 12% | 19% |

#### Cumulative incidence of myocardial infarction, accounting for competing risk of death

Ischaemic stroke

| | 6 months | 1 year | 5 years | 10 years |
|---|---|---|---|---|
| Male, ≤ 70 years | 0.79% | 1.2% | 3.7% | 6.5% |
| Male, > 70 | 1.3% | 1.9% | 5.2% | 6.8% |
| Female, ≤ 70 years | 0.75% | 0.98% | 2.6% | 4.7% |
| Female, > 70 years | 1.2% | 1.6% | 4.0% | 5.4% |

Intracerebral haemorrhage

| | 6 months | 1 year | 5 years | 10 years |
|---|---|---|---|---|
| Male, ≤ 70 years | 0.75% | 1.1% | 2.2% | 5.1% |
| Male, > 70 | 0.61% | 1.1% | 2.7% | 3.6% |
| Female, ≤ 70 years | 0.21% | 0.21% | 0.86% | 1.5% |
| Female, > 70 years | 0.35% | 0.67% | 1.8% | 2.4% |

Subarachnoid haemorrhage

| | 6 months | 1 year | 5 years | 10 years |
|---|---|---|---|---|
| Male, ≤ 70 years | 0.14% | 0.55% | 1.8% | 3.9% |
| Male, > 70 | 1.1% | 1.1% | 1.9% | 2.5% |
| Female, ≤ 70 years | 0.00% | 0.14% | 0.97% | 1.9% |
| Female, > 70 years | 0.17% | 0.50% | 1.9% | 2.6% |

Subdural haemorrhage

| | 6 months | 1 year | 5 years | 10 years |
|---|---|---|---|---|
| Male, ≤ 70 years | 0.00% | 0.00% | 1.7% | 5.3% |
| Male, > 70 | 1.4% | 1.7% | 4.1% | 6.5% |
| Female, ≤ 70 years | 0.00% | 0.00% | 0.86% | 2.4% |
| Female, > 70 years | 1.4% | 1.7% | 3.4% | 3.9% |

eFigure 4 Heatmaps of cumulative incidence of outcomes for stroke subtypes, stratified by age and sex.

#### Cumulative incidence of cancer, accounting for competing risk of death

Ischaemic stroke, cortical

| | 6 months | 1 year | 5 years | 10 years |
|---|---|---|---|---|
| Male, ≤ 70 years | 1.4% | 2.2% | 6.2% | 10% |
| Male, > 70 | 1.7% | 2.6% | 7.0% | 11% |
| Female, ≤ 70 years | 1.4% | 1.9% | 5.5% | 8.5% |
| Female, > 70 years | 1.2% | 1.8% | 4.6% | 7.2% |

Ischaemic stroke, deep

| | 6 months | 1 year | 5 years | 10 years |
|---|---|---|---|---|
| Male, ≤ 70 years | 0.91% | 1.6% | 6.3% | 13% |
| Male, > 70 | 1.6% | 2.8% | 9.8% | 14% |
| Female, ≤ 70 years | 0.98% | 1.7% | 6.1% | 11% |
| Female, > 70 years | 0.91% | 1.8% | 5.9% | 8.5% |

Intracerebral haemorrhage, lobar

| | 6 months | 1 year | 5 years | 10 years |
|---|---|---|---|---|
| Male, ≤ 70 years | 1.4% | 1.4% | 3.3% | 4.2% |
| Male, > 70 | 1.6% | 2.3% | 6.2% | 7.5% |
| Female, ≤ 70 years | 0.00% | 0.59% | 3.3% | 3.3% |
| Female, > 70 years | 0.95% | 1.9% | 3.6% | 5.0% |

Intracerebral haemorrhage, deep

| | 6 months | 1 year | 5 years | 10 years |
|---|---|---|---|---|
| Male, ≤ 70 years | 1.6% | 1.8% | 4.8% | 6.7% |
| Male, > 70 | 1.3% | 2.5% | 5.5% | 9.0% |
| Female, ≤ 70 years | 0.45% | 0.45% | 2.9% | 9.8% |
| Female, > 70 years | 0.59% | 1.5% | 3.7% | 5.7% |

#### Cumulative incidence of readmission with stroke, accounting for competing risk of death

Ischaemic stroke, cortical

| | 6 months | 1 year | 5 years | 10 years |
|---|---|---|---|---|
| Male, ≤ 70 years | 3.4% | 4.2% | 8.4% | 12% |
| Male, > 70 | 3.2% | 4.3% | 8.8% | 11% |
| Female, ≤ 70 years | 2.8% | 3.9% | 8.0% | 12% |
| Female, > 70 years | 3.1% | 4.2% | 7.9% | 9.6% |

Ischaemic stroke, deep

| | 6 months | 1 year | 5 years | 10 years |
|---|---|---|---|---|
| Male, ≤ 70 years | 3.2% | 4.2% | 9.4% | 13% |
| Male, > 70 | 2.9% | 3.9% | 8.8% | 11% |
| Female, ≤ 70 years | 3.6% | 4.5% | 9.6% | 14% |
| Female, > 70 years | 2.8% | 4.0% | 8.3% | 10% |

Intracerebral haemorrhage, lobar

| | 6 months | 1 year | 5 years | 10 years |
|---|---|---|---|---|
| Male, ≤ 70 years | 5.0% | 5.5% | 8.8% | 15% |
| Male, > 70 | 4.6% | 5.5% | 11% | 13% |
| Female, ≤ 70 years | 4.7% | 4.7% | 10% | 17% |
| Female, > 70 years | 2.5% | 4.4% | 8.9% | 10% |

Intracerebral haemorrhage, deep

| | 6 months | 1 year | 5 years | 10 years |
|---|---|---|---|---|
| Male, ≤ 70 years | 3.5% | 4.2% | 8.5% | 10% |
| Male, > 70 | 1.9% | 2.5% | 8.2% | 9.9% |
| Female, ≤ 70 years | 1.4% | 2.7% | 8.0% | 10% |
| Female, > 70 years | 2.6% | 3.7% | 6.3% | 8.2% |

Cumulative incidence (%): 0, 10, 20, 30

#### Cumulative incidence of epilepsy, accounting for competing risk of death

Ischaemic stroke, cortical

| | 6 months | 1 year | 5 years | 10 years |
|---|---|---|---|---|
| Male, ≤ 70 years | 4.8% | 7.4% | 13% | 14% |
| Male, > 70 | 2.9% | 4.3% | 6.8% | 7.4% |
| Female, ≤ 70 years | 4.4% | 6.3% | 11% | 14% |
| Female, > 70 years | 2.4% | 3.9% | 6.2% | 7.1% |

Ischaemic stroke, deep

| | 6 months | 1 year | 5 years | 10 years |
|---|---|---|---|---|
| Male, ≤ 70 years | 2.3% | 3.4% | 5.2% | 6.0% |
| Male, > 70 | 2.1% | 2.7% | 4.4% | 5.2% |
| Female, ≤ 70 years | 2.8% | 3.6% | 6.2% | 8.7% |
| Female, > 70 years | 2.3% | 3.0% | 4.9% | 5.2% |

Intracerebral haemorrhage, lobar

| | 6 months | 1 year | 5 years | 10 years |
|---|---|---|---|---|
| Male, ≤ 70 years | 13% | 14% | 18% | 20% |
| Male, > 70 | 4.5% | 6.2% | 9.9% | 11% |
| Female, ≤ 70 years | 10% | 14% | 20% | 20% |
| Female, > 70 years | 4.2% | 6.4% | 8.5% | 9.1% |

Intracerebral haemorrhage, deep

| | 6 months | 1 year | 5 years | 10 years |
|---|---|---|---|---|
| Male, ≤ 70 years | 3.2% | 4.1% | 6.7% | 8.5% |
| Male, > 70 | 1.3% | 2.1% | 3.0% | 4.2% |
| Female, ≤ 70 years | 3.3% | 6.2% | 8.7% | 11% |
| Female, > 70 years | 2.2% | 2.2% | 3.0% | 3.0% |

#### Cumulative incidence of myocardial infarction, accounting for competing risk of death

Ischaemic stroke, cortical

| | 6 months | 1 year | 5 years | 10 years |
|---|---|---|---|---|
| Male, ≤ 70 years | 0.97% | 1.3% | 3.9% | 6.6% |
| Male, > 70 | 1.6% | 2.3% | 5.4% | 6.9% |
| Female, ≤ 70 years | 0.86% | 0.99% | 3.0% | 5.3% |
| Female, > 70 years | 1.3% | 1.7% | 3.8% | 5.1% |

Ischaemic stroke, deep

| | 6 months | 1 year | 5 years | 10 years |
|---|---|---|---|---|
| Male, ≤ 70 years | 0.84% | 1.3% | 4.5% | 6.9% |
| Male, > 70 | 1.2% | 1.6% | 5.7% | 7.3% |
| Female, ≤ 70 years | 0.28% | 0.56% | 2.2% | 4.8% |
| Female, > 70 years | 1.3% | 1.8% | 4.7% | 6.2% |

Intracerebral haemorrhage, lobar

| | 6 months | 1 year | 5 years | 10 years |
|---|---|---|---|---|
| Male, ≤ 70 years | 1.4% | 1.4% | 2.0% | 5.5% |
| Male, > 70 | 0.69% | 1.4% | 2.8% | 3.4% |
| Female, ≤ 70 years | 0.00% | 0.00% | 2.4% | 2.4% |
| Female, > 70 years | 0.00% | 0.32% | 1.6% | 2.3% |

Intracerebral haemorrhage, deep

| | 6 months | 1 year | 5 years | 10 years |
|---|---|---|---|---|
| Male, ≤ 70 years | 0.92% | 1.2% | 2.4% | 5.3% |
| Male, > 70 | 0.63% | 0.84% | 3.6% | 5.4% |
| Female, ≤ 70 years | 0.00% | 0.00% | 0.00% | 0.73% |
| Female, > 70 years | 0.15% | 0.44% | 2.1% | 2.3% |

eTable 4: Causes of death for people with each stroke type and the combined control population

| *Ranking* | **Ischemic stroke** | **ICH** | **SAH** | **SDH** | **Control** |
|---|---|---|---|---|---|
| *First* | I64 Stroke, not specified as hemorrhage or infarction (13.3%) | I61 Intracerebral hemorrhage (44.7%) | I60 Subarachnoid hemorrhage (52.9%) | I62 Other nontraumatic intracranial hemorrhage (26.1%) | I21 Acute myocardial infarction (7.2%) |
| *Second* | I63 Cerebral infarction (13.3%) | I62 Other nontraumatic intracranial hemorrhage (9%) | I61 Intracerebral hemorrhage (7.2%) | W19 Unspecified fall (4.9%) | F03 Unspecified dementia (5.8%) |
| *Third* | I69 Sequelae of cerebrovascular disease (8.9%) | I69 Sequelae of cerebrovascular disease (6.4%) | I62 Other nontraumatic intracranial hemorrhage (5.5%) | F01 Vascular dementia (3.7%) | G30 Alzheimer disease (5.4%) |

### eFigure 5. Forest plot for death, stratified by sex and age

| Outcome | Stroke type | Hazard ratio | N with outcome |
|---|---|---|---|
| Death before 6 months | Ischaemic stroke, Male | 7.5 (6.97–8.08) | 4671 |
| | Intracerebral haemorrhage, Male | 23.57 (20.18–27.55) | 1435 |
| | Subarachnoid haemorrhage, Male | 34.71 (23.41–51.46) | 286 |
| | Subdural haemorrhage, Male | 9.54 (7.53–12.08) | 522 |
| | Ischaemic stroke, Female | 9.58 (8.99–10.21) | 6467 |
| | Intracerebral haemorrhage, Female | 22.86 (19.9–26.27) | 1855 |
| | Subarachnoid haemorrhage, Female | 45.02 (33.83–59.91) | 590 |
| | Subdural haemorrhage, Female | 12.5 (9.46–16.52) | 375 |
| | Ischaemic stroke, Less than or equal to 70 years | 15.63 (13.37–18.28) | 1534 |
| | Intracerebral haemorrhage, Less than or equal to 70 years | 78.04 (56.15–108.46) | 755 |
| | Subarachnoid haemorrhage, Less than or equal to 70 years | 77.46 (49.47–121.29) | 414 |
| | Subdural haemorrhage, Less than or equal to 70 years | 30.32 (15.4–59.67) | 134 |
| | Ischaemic stroke, Above 70 years | 7.23 (6.87–7.61) | 9604 |
| | Intracerebral haemorrhage, Above 70 years | 18.26 (16.3–20.45) | 2535 |
| | Subarachnoid haemorrhage, Above 70 years | 30.23 (22.76–40.16) | 462 |
| | Subdural haemorrhage, Above 70 years | 8.63 (7.14–10.44) | 763 |
| Death beyond 6 months | Ischaemic stroke, Male | 2.27 (2.19–2.36) | 12798 |
| | Intracerebral haemorrhage, Male | 3.91 (3.55–4.3) | 2191 |
| | Subarachnoid haemorrhage, Male | 4.02 (3.17–5.09) | 394 |
| | Subdural haemorrhage, Male | 2.05 (1.79–2.35) | 1057 |
| | Ischaemic stroke, Female | 2.51 (2.42–2.6) | 15154 |
| | Intracerebral haemorrhage, Female | 4.43 (4.07–4.82) | 2823 |
| | Subarachnoid haemorrhage, Female | 4.47 (3.79–5.27) | 787 |
| | Subdural haemorrhage, Female | 2.79 (2.36–3.29) | 710 |
| | Ischaemic stroke, Less than or equal to 70 years | 2.06 (1.95–2.17) | 5797 |
| | Intracerebral haemorrhage, Less than or equal to 70 years | 2.53 (2.14–2.99) | 637 |
| | Subarachnoid haemorrhage, Less than or equal to 70 years | 2 (1.23–3.25) | 67 |
| | Subdural haemorrhage, Less than or equal to 70 years | 1.8 (1.43–2.27) | 350 |
| | Ischaemic stroke, Above 70 years | 1.57 (1.51–1.63) | 11017 |
| | Intracerebral haemorrhage, Above 70 years | 1.99 (1.75–2.27) | 1087 |
| | Subarachnoid haemorrhage, Above 70 years | 1.58 (1.21–2.06) | 238 |
| | Subdural haemorrhage, Above 70 years | 1.35 (1.12–1.63) | 520 |

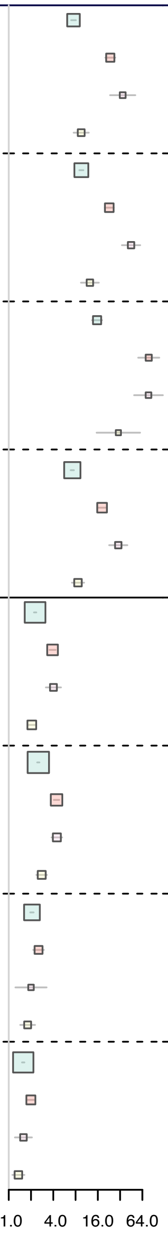

eFigure 6. Forest plots for all outcomes by stroke subtype, including those with no location.

A. Early after stroke (<6 months)

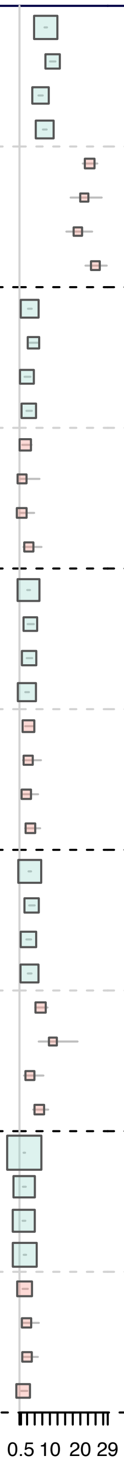

| Outcome | Stroke type | Hazard ratio | N with outcome |
|---|---|---|---|
| Death | Ischaemic stroke | 8.63 (8.22–9.05) | 11138 |
| | Ischaemic stroke, Cortical | 10.89 (10.02–11.83) | 4134 |
| | Ischaemic stroke, Deep | 6.91 (6.24–7.66) | 2202 |
| | Ischaemic stroke, No location | 8.29 (7.7–8.94) | 4595 |
| | Intracerebral haemorrhage | 23.06 (20.79–25.58) | 3290 |
| | Intracerebral haemorrhage, Lobar | 21.3 (16.8–27.01) | 626 |
| | Intracerebral haemorrhage, Deep | 19.18 (15.41–23.88) | 706 |
| | Intracerebral haemorrhage, No location | 24.93 (21.68–28.66) | 1880 |
| MI | Ischaemic stroke | 3.35 (2.79–4.03) | 568 |
| | Ischaemic stroke, Cortical | 4.6 (3.35–6.31) | 189 |
| | Ischaemic stroke, Deep | 2.45 (1.66–3.61) | 135 |
| | Ischaemic stroke, No location | 3.06 (2.29–4.08) | 235 |
| | Intracerebral haemorrhage | 1.91 (0.92–3.96) | 35 |
| | Intracerebral haemorrhage, Lobar | 0.85 (0.11–6.57) | <10 |
| | Intracerebral haemorrhage, Deep | 0.73 (0.11–4.83) | <10 |
| | Intracerebral haemorrhage, No location | 3.07 (1.31–7.23) | 21 |
| Dementia | Ischaemic stroke | 2.97 (2.55–3.46) | 750 |
| | Ischaemic stroke, Cortical | 3.57 (2.7–4.71) | 234 |
| | Ischaemic stroke, Deep | 3.16 (2.38–4.19) | 208 |
| | Ischaemic stroke, No location | 2.44 (1.91–3.12) | 294 |
| | Intracerebral haemorrhage | 2.95 (1.87–4.66) | 99 |
| | Intracerebral haemorrhage, Lobar | 2.86 (1.15–7.07) | 29 |
| | Intracerebral haemorrhage, Deep | 2.24 (0.82–6.13) | 18 |
| | Intracerebral haemorrhage, No location | 3.59 (1.89–6.81) | 50 |
| Epilepsy | Ischaemic stroke | 3.37 (2.97–3.82) | 1385 |
| | Ischaemic stroke, Cortical | 3.99 (3.18–5) | 436 |
| | Ischaemic stroke, Deep | 2.94 (2.26–3.84) | 286 |
| | Ischaemic stroke, No location | 3.29 (2.73–3.96) | 649 |
| | Intracerebral haemorrhage | 6.94 (5.23–9.22) | 310 |
| | Intracerebral haemorrhage, Lobar | 10.98 (6.32–19.08) | 87 |
| | Intracerebral haemorrhage, Deep | 3.43 (1.49–7.86) | 41 |
| | Intracerebral haemorrhage, No location | 6.51 (4.49–9.45) | 177 |
| Cancer | Ischaemic stroke | 1.58 (1.35–1.85) | 716 |
| | Ischaemic stroke, Cortical | 1.54 (1.14–2.08) | 191 |
| | Ischaemic stroke, Deep | 1.41 (1.03–1.93) | 184 |
| | Ischaemic stroke, No location | 1.74 (1.39–2.19) | 330 |
| | Intracerebral haemorrhage | 1.68 (1.03–2.75) | 70 |
| | Intracerebral haemorrhage, Lobar | 2.32 (0.85–6.37) | 16 |
| | Intracerebral haemorrhage, Deep | 2.48 (1.01–6.05) | 23 |
| | Intracerebral haemorrhage, No location | 1.21 (0.57–2.58) | 29 |

#### B. Late after stroke (>6 months)

| Outcome | Stroke type | Hazard ratio | N with outcome |
|---|---|---|---|
| Death | Ischaemic stroke | 1.88 (1.82–1.94) | 16814 |
| | Ischaemic stroke, Cortical | 2.04 (1.92–2.17) | 4879 |
| | Ischaemic stroke, Deep | 1.86 (1.75–1.97) | 4616 |
| | Ischaemic stroke, No location | 1.82 (1.73–1.91) | 7098 |
| | Intracerebral haemorrhage | 2.36 (2.13–2.61) | 1724 |
| | Intracerebral haemorrhage, Lobar | 2.58 (2.07–3.22) | 407 |
| | Intracerebral haemorrhage, Deep | 2.32 (1.9–2.83) | 467 |
| | Intracerebral haemorrhage, No location | 2.41 (2.08–2.78) | 814 |
| MI | Ischaemic stroke | 1.34 (1.22–1.48) | 1777 |
| | Ischaemic stroke, Cortical | 1.16 (0.96–1.41) | 476 |
| | Ischaemic stroke, Deep | 1.65 (1.39–1.96) | 510 |
| | Ischaemic stroke, No location | 1.29 (1.12–1.49) | 765 |
| | Intracerebral haemorrhage | 1.49 (1.07–2.07) | 138 |
| | Intracerebral haemorrhage, Lobar | 1.51 (0.72–3.17) | 29 |
| | Intracerebral haemorrhage, Deep | 1.77 (0.97–3.24) | 42 |
| | Intracerebral haemorrhage, No location | 1.46 (0.91–2.34) | 65 |
| Dementia | Ischaemic stroke | 1.97 (1.84–2.11) | 3578 |
| | Ischaemic stroke, Cortical | 2.16 (1.9–2.46) | 1057 |
| | Ischaemic stroke, Deep | 1.94 (1.71–2.21) | 971 |
| | Ischaemic stroke, No location | 1.94 (1.75–2.15) | 1504 |
| | Intracerebral haemorrhage | 2.54 (2.05–3.15) | 407 |
| | Intracerebral haemorrhage, Lobar | 3.49 (2.3–5.29) | 124 |
| | Intracerebral haemorrhage, Deep | 2.27 (1.47–3.49) | 90 |
| | Intracerebral haemorrhage, No location | 2.42 (1.77–3.31) | 185 |
| Epilepsy | Ischaemic stroke | 3.63 (3.27–4.02) | 1946 |
| | Ischaemic stroke, Cortical | 5.62 (4.68–6.75) | 709 |
| | Ischaemic stroke, Deep | 2.35 (1.87–2.95) | 354 |
| | Ischaemic stroke, No location | 3.59 (3.08–4.18) | 862 |
| | Intracerebral haemorrhage | 8.24 (6.2–10.96) | 279 |
| | Intracerebral haemorrhage, Lobar | 9.33 (5.29–16.48) | 77 |
| | Intracerebral haemorrhage, Deep | 4.7 (2.33–9.48) | 43 |
| | Intracerebral haemorrhage, No location | 9.83 (6.65–14.54) | 152 |
| Cancer | Ischaemic stroke | 1.07 (1–1.14) | 3948 |
| | Ischaemic stroke, Cortical | 1.03 (0.91–1.18) | 887 |
| | Ischaemic stroke, Deep | 1.11 (0.98–1.24) | 1068 |
| | Ischaemic stroke, No location | 1.08 (0.99–1.18) | 1946 |
| | Intracerebral haemorrhage | 0.88 (0.7–1.1) | 318 |
| | Intracerebral haemorrhage, Lobar | 0.67 (0.39–1.17) | 61 |
| | Intracerebral haemorrhage, Deep | 1.26 (0.83–1.89) | 88 |
| | Intracerebral haemorrhage, No location | 0.77 (0.56–1.06) | 163 |

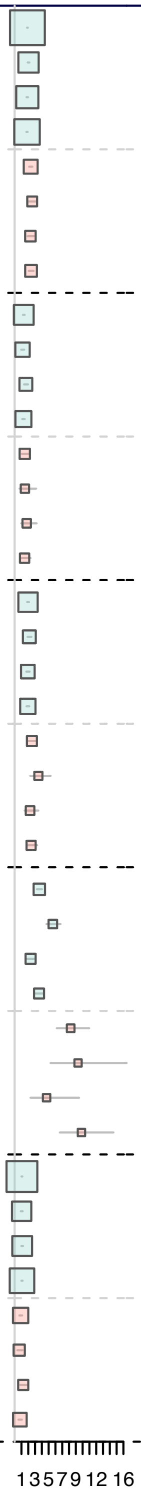
